# Development and Initial Validation of the Duke Misophonia Questionnaire

**DOI:** 10.1101/2021.05.05.21256694

**Authors:** M. Zachary Rosenthal, Deepika Anand, Clair Robbins, Zachary J. Williams, Rachel Guetta, Jacqueline Trumbull, Lisalynn Kelly

## Abstract

Misophonia is characterized by decreased tolerance and accompanying defensive motivational system responding to certain aversive sounds and contextual cues associated with such stimuli, typically repetitive oral (e.g., eating sounds) or nasal (e.g., breathing sounds) stimuli. Responses elicit significant psychological distress and impairment in functioning, and include acute increases in (a) negative affect (e.g., anger, anxiety, and disgust), (b) physiological arousal (e.g., sympathetic nervous system activation), and (c) overt behavior (e.g., escape behavior and verbal aggression toward individuals generating triggers). A major barrier to research and treatment of misophonia is the lack of rigorously validated assessment measures. As such, the primary purpose of this study was to develop and psychometrically validate a self-report measure of misophonia, the Duke Misophonia Questionnaire (DMQ). There were two phases of measure development. In Phase 1, items were generated and iteratively refined from a combination of the scientific literature and qualitative feedback from misophonia sufferers, their family members, and professional experts. In Phase 2, a large community sample of adults (*n* = 424) completed DMQ candidate items and other measures needed for psychometric analyses. A series of iterative analytic procedures (e.g., factor analyses and IRT) were used to derive final DMQ items and scales. The final DMQ is 86 items, and includes subscales: (1) Trigger frequency (16 items), (2) Affective Responses (5 items), (3) Physiological Responses (8 items), (4) Cognitive Responses (10 items), (5) Coping Before (6 items), (6) Coping During (10 items), (7) Coping After (5 items), (8) Impairment (12 items), and Beliefs (14 items). Composite scales were derived for overall Symptom Severity (combined Affective, Physiological, and Cognitive subscales) and Coping (combined the three Coping subscales). The analytic procedures used enable administration of the total DMQ, individual subscales, or the derived composite scales.

---

Originally described by Jastreboff and Jastreboff in 2001, misophonia is characterized by decreased tolerance and accompanying defensive motivational system responding to certain aversive sounds and contextual cues associated with such stimuli (Brout et al., 2018). Cues (sometimes called “triggers”) commonly are repetitive oral (e.g., eating, chewing, throat clearing) or nasal (e.g., heavy breathing, sniffing; Jager et al., 2020) stimuli. These triggering cues typically are produced by other humans but can be animal-produced (e.g., pets grooming themselves) or generated environmentally (e.g., clock ticking). Responses elicited by triggering stimuli may be experienced by those with misophonia as highly unpleasant, and can elicit sudden changes in multi-modal processes underlying emotional functioning and associated sequelae, including (a) subjective affective state (e.g., increases in irritation, anger, anxiety, and disgust), (b) physiological arousal (e.g., sympathetic nervous system activation), and (c) overt behavior (e.g., escape behavior and verbal aggression toward individuals generating triggers; e.g., Brout et al., 2018; Edelstein et al., 2014; Rouw & Erfanian, 2018).

Although the prevalence of misophonia has not been rigorously investigated, studies with university students in the United States, China, and the United Kingdom have found between 12% and 20% of respondents experience moderate or higher symptoms of misophonia causing significant distress and impairment in daily functioning (Naylor, Camaino, Scutt, Hoare, and Baguley, 2020; Wu, Lewin, Murphy, & Storch, 2014; Zhou, Wu, & Storch, 2017). Higher symptoms of misophonia are associated with greater psychological distress and a range of transdiagnostic vulnerability factors for psychopathology, such as anxiety symptoms (e.g., McKay et al., 2018; Quek et al., 2018; Wu et al., 2014). Additionally, trait neuroticism, perfectionism, and difficulties with emotion regulation are associated with misophonia (Cassiello-Robbins et al., 2020; Jager et al., 2020). The association between greater misophonia symptoms and problems with mental health has been observed in studies across many countries, including China, Singapore, Brazil, Poland, the Netherlands, the United Kingdom, and the United States. Although little is known about the influence of cultural factors on the etiology, presentation, or maintenance of misophonia, the available scientific evidence indicates it is not a phenomenon limited to a specific country or culture. Additionally, although some studies have sampled participants from online misophonia support forums (e.g., Rouw & Erfanian, 2018), other studies have recruited university students (Wu et al., 2014; Zhou et al., 2017), medical students (Naylor et al., 2020), community samples (e.g., Cassiello-Robbins et al., 2020; Cassiello-Robbins et al., 2021), and individuals seeking treatment (Jager et al., 2020; Quek et al., 2018; Schröder et al., 2013). The diversity of sampling approaches suggests that misophonia can be studied in a general population, and is not limited to those with access to online and social media platforms dedicated to this condition.

Research examining the relationship between misophonia and psychiatric disorders has recently begun. Individuals with misophonia report having current symptoms of a range of co-occurring psychiatric conditions, including but not limited to anxiety, mood, and personality disorders (Cassiello-Robbins et al., 2020; Claiborn et al., 2020; Jager et al., 2020; Rouw & Erfanian, 2018). Although early studies on this topic led to speculations that misophonia could be considered an obsessive-compulsive psychiatric disorder (Schröder et al., 2013), the accruing empirical data indicates otherwise. Indeed, the available published scientific research suggests that misophonia does not appear to be uniquely associated with any specific psychiatric disorder or class of disorders (Jager et al., 2020). The most consistent findings regarding co-occurrence suggest the possibility that misophonia may be likely to co-occur with symptoms of anxiety, mood, and personality disorders, but not with any one single diagnosis (Cassiello-Robbins et al., 2021; Jager et al., 2020). More specifically, some studies have found that misophonia may be most highly correlated with anxiety symptoms (e.g., Siepsak, Sobczak, Bohaterewicz, Cichocki, & Dragan, 2020) or anxiety disorders (e.g., Cassiello-Robbins et al., 2021), and that anxiety significantly mediates the relationship between misophonia symptoms and rage (Wu et al., 2014). This is congruent with research indicating that sensory over-responsivity during childhood is a vulnerability factor associated with the development of anxiety during adulthood (Carpenter et al., 2019; McMahon, Anand, Morris-Jones, & Rosenthal, 2019).

As a caveat, it should be noted that with few exceptions (Jager et al., 2020), most studies reporting about the relationship between misophonia and psychiatric disorders have had very small samples (e.g., Cassiello Robbins et al., 2021). Others with large samples have used self-report questionnaires (Rouw & Erfanian, 2018), asking respondents to indicate if they have ever been diagnosed with certain selected psychiatric disorders. Because not all disorders in these studies were included as response options, and individuals do not always know which specific diagnoses they have been given by mental health professionals, studies using self-report as a method for assessing psychiatric diagnosis yield inadequately limited inferences. Until additional rigorous research is done using structured and psychometrically validated psychiatric diagnostic interviews (e.g., see Jager et al., 2020 for the first such study), it is premature to arrive at definitive conclusions about the relationship between misophonia and psychiatric disorders.

Individuals with misophonia may present for clinical services to providers across a range of professional disciplines, including but not limited to primary care physicians, pediatricians, occupational therapists, audiologists, and those providing mental health services. Accordingly, psychometrically validated assessment measures of misophonia are needed for use in these clinical settings. As clinical observations and the first studies of misophonia were conducted, a number of self-report inventories were developed as a way to preliminarily quantify severity of misophonia and associated impairments in functioning. However, there are important weaknesses in most of these early measures. Chief limitations include: (a) reliance on expert opinion to generate items, without including a wider range of stakeholders such as sufferers and their family members, (b) restricting the item pool to assessment of symptoms and functional impairment, thereby not including items reflecting underlying mechanistic processes that could be targeted for change in treatment (e.g., difficulties with the regulation of attention, emotion, behavior, physiological arousal, and cognition before, during, or after being triggered), (c) item pools were not subjected to iterative psychometric analyses to derive best fitting models, and (d) final items and subscales of many of these measures were not subjected to psychometric validation. The purpose of the present study was to develop and provide initial psychometric validation of a new measure of misophonia that helps to address the limitations of extent measures.

## A Review of Extant Self-Report Measures of Misophonia

One prominent measure of misophonia is the Misophonia Questionnaire (MQ; Wu et al., 2014). Developed by experts based on a literature review and their own clinical experience, this self-report measure consists of three subscales using a Likert-type scale: (a) frequency of specific trigger sounds, (b) frequency of certain emotions and behavioral responses to trigger sounds, and (c) overall perception of severity of sound sensitivities. This last subscale is a single item ranging from 1 to 15 that asks the participant to indicate how severe the impact of their sound sensitivity is on their life, with higher scores indicating greater impact. The authors suggest a score above 6 indicates clinically significant symptoms. In the initial validation paper, which utilized a college student sample, the authors reported good internal consistency for the first two subscales (.86 for each and .89 for total score). The authors also noted preliminary convergent validity with the Adult Sensory Questionnaire (ASQ; Kinnealey & Oliver, 2002; Kinnealey, Oliver, and Willbarger, 1995), a measure of multi-sensory responsivity (*r* = .50). Finally, they indicated preliminary discriminant validity as the correlation between MQ total score and ASQ “sound sensitivities” subscale was significantly different from the correlation with responsivity in other sensory modalities. Although the initial psychometrics for the MQ are promising, the use of a university sample renders its generalizability unclear to the broader population of misophonia sufferers. Additionally, the use of a single self-reported severity item limits sufficient psychometric evaluation. Despite these limitations, the MQ is among the most frequently used measure of misophonia (Cassiello-Robbins et al., 2020; Daniels et al., 2020; Frank & McKay, 2019; Frank et al., 2020; McKay et al., 2018; Zhou et al., 2017) and has been influential in the early scientific findings in this field.

The Amsterdam Misophonia Scale (A-MISO-S; Schröder et al., 2013) is another commonly used self-report measure. Adapted from the Yale-Brown Obsessive-Compulsive Scale (Goodman et al., 1989) to reflect items consistent with the assessment of obsessive compulsive disorder (OCD), the A-MISO-S consists of 6 items using a Likert-type scale that assess, during the past week: time occupied by misophonic sounds, interference with daily functioning caused by trigger sounds, distress caused by misophonic sounds, efforts to resist thoughts about trigger sounds, control over thoughts about misophonic sounds, and avoidance caused by misophonia. A final item is used for free responses and assesses the worst feared consequence of not being able to avoid misophonic triggers. The A-MISO-S was preliminarily validated in a sample of medical students and demonstrated good internal consistency (α = .81; Naylor et al., 2020). As with the MQ, additional studies are needed with the A-MISO-S using community samples in order to enhance the generalizability of this measure. Although the items selected for the A-MISO-S were created by adapting a measure of OCD, accruing scientific data suggests that misophonia may not be significantly associated with OCD relative to other psychiatric disorders (Jager et al., 2020). Additionally, one study using a large community sample of adults reported that misophonia is negatively correlated with some features of OCD, and positively correlated with other features, suggesting a complex relationship between misophonia and OCD (McKay, Kim, Mancusi, Storch, & Spankovich, 2018). A related limitation to the A-MISO-S is that item generation and refinement was not done using psychometric analyses. Nonetheless, the original A-MISO-S and a revised version have been widely used in misophonia research across multiple countries, suggesting it merits additional psychometric evaluation and is promising as a useful and brief measure (Eijsker et al., 2019; Erfanian et al., 2018, 2019a, 2019b; Jager et al., 2020; Kluckow et al., 2014; Natalini et al., 2020; Naylor et al., 2020; Quek et al., 2018; Rouw et al., 2017; Schröder et al., 2014).

The MisoQuest (Siepsiak, Silwerski, & Dragan, 2020) is the most recently developed measure of misophonia. This self-report measure has undergone the most extensive psychometric evaluation of any misophonia self-report inventory to date. From a pool of 60 items, created based on proposed criteria for a diagnosis of misophonia (Schröder et al., 2013), exploratory factor analysis, confirmatory factor analysis, and item response theory (IRT) analyses derived 14 items that comprise the final scale. Scores range from 0–70, with higher scores indicating greater misophonia severity, and the authors suggest a score of 61 be used for clinical cut off. Initial reports suggest good reliability (.96) as well as test-retest reliability (.84). Although this measure is psychometrically robust, the items were generated using proposed diagnostic criteria that have not been empirically validated. Different diagnostic criteria for misophonia have been proposed (Dozier, Lopez, & Pearson, 2017; Schröder et al., 2013) and the nature and boundaries around misophonia are scientifically unknown (Taylor, 2017). As a result, there are no empirically derived diagnostic criteria. Indeed, an expert consensus definition of misophonia has only recently been completed (Swedo et al., 2021). Because the MisoQuest was limited to items based on one proposed set of diagnostic criteria and did not including a wider range of sources for item generation (e.g., other proposed models; other stakeholders, such as sufferers and their loved ones), it is unclear to what extent this measure adequately taps the broader construct of misophonia. In addition, this measure was written and validated with a Polish sample, rendering it unknown whether the MisoQuest is a reliable and valid measure for individuals with English as a first language. Because it was developed with rigorous attention to psychometric validation, the MisoQuest is a measure warranting further cross-validation in English-speaking and other diverse samples.

The Selective Sound Sensitivity Syndrome Scale (S-Five; Vitoratou et al., 2020) is new self-report measure under development. This scale was developed by the authors and has undergone three waves of data collection and psychometric evaluation using a community sample in the United Kingdom. The S-Five consists of five factors: internalizing appraisals, externalizing appraisals, perceived threat and avoidance behavior, outbursts, and impact on functioning. The pre-print manuscript indicates detailed psychometric data will be presented in a peer reviewed journal article currently under preparation.

A number of other measures of misophonia exist, including the Misophonia Coping Responses Survey (MCRS; Dozier, 2015), Misophonia Activation Scale (MAS-1; http://www.misophonia-uk.org/the-misophonia-activation-scale.html), Misophonia Emotion Responses (MER-2; Dozier, 2015), Misophonia Physiological Responses Scale (MPRS; Bauman & Dozier), and Misophonia Trigger Severity Scale (Dozier, 2015). These measures have yet to undergo psychometric evaluation and have been used somewhat less frequently in the published research literature.

Beyond the weaknesses of individual extant measures, these instruments typically focus on symptoms and impairment. No measure to date has looked comprehensively at affective, cognitive, behavioral, and physiological processes that occur in response to misophonic cues, as well as efforts to cope with misophonia or how misophonia affects one’s self-concept. However, it is important to assess this condition holistically in order to develop targeted interventions (i.e., individuals who report problematic cognitions as the most distressing or impairing feature of misophonia may benefit from a different intervention than those who report primary problems of hypervigilance or excessive avoidance behavior) and provide a comprehensive understanding of an individual’s experience of misophonia.

## Current Study

The overarching aim of the present study was to develop and conduct initial validation of a new self-report measure of misophonia for adults. The *a priori* goals were to identify and psychometrically evaluate items representing one’s current: (a) frequency of being affected by misophonic sounds, (b) degree of responding to misophonic sounds across domains of functioning (i.e., affective, physiological, cognitive and behavioral responses to trigger cues), (c) social and occupational/academic impairment caused by misophonia, (d) ways of coping with misophonic cues before, during, and after the onset of these stimuli, and (e) consistent with the cognitive theory for emotional disorders (Beck, 1996), beliefs about self, others, and the world related to the experience of misophonia symptoms. The new measure was developed with the intention of being used in scientific and clinical settings to comprehensively characterize the severity, adverse impact, and ways of coping with of misophonia.

## Method

The Duke Misophonia Questionnaire (DMQ) was developed in two phases. In Phase 1, the item pool was developed using a literature review, stakeholder responses to a qualitative interview (sufferers and loved ones of those with misophonia), and iterative feedback from expert professionals and sufferers. Phase 2 involved rigorous psychometric refinement and validation. A summary of the methodology used to develop the DMQ is depicted in Figures 1a-1c. Details of each element in this flowchart are described below.

**Figure 1a.**
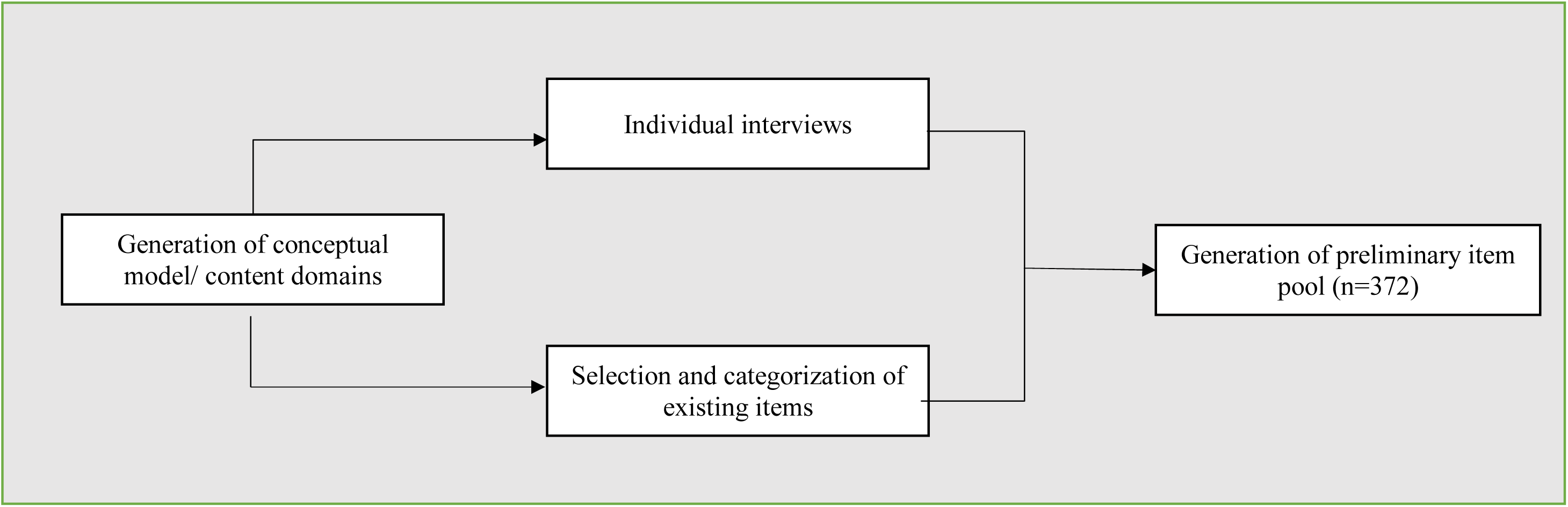
Development of Item Pool.

**Figure 1b.**
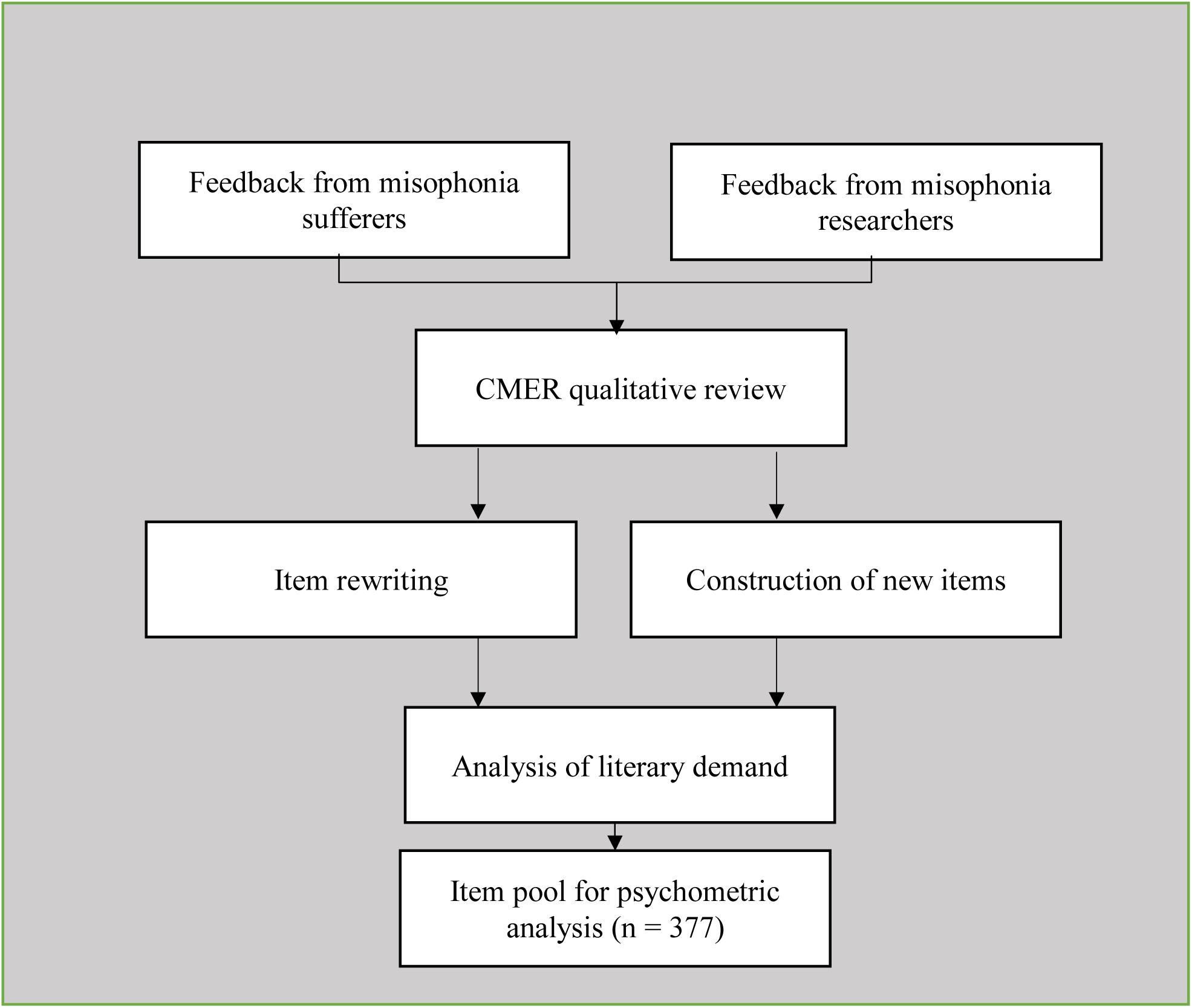
Refinement of Item Pool.

**Figure 1c.**
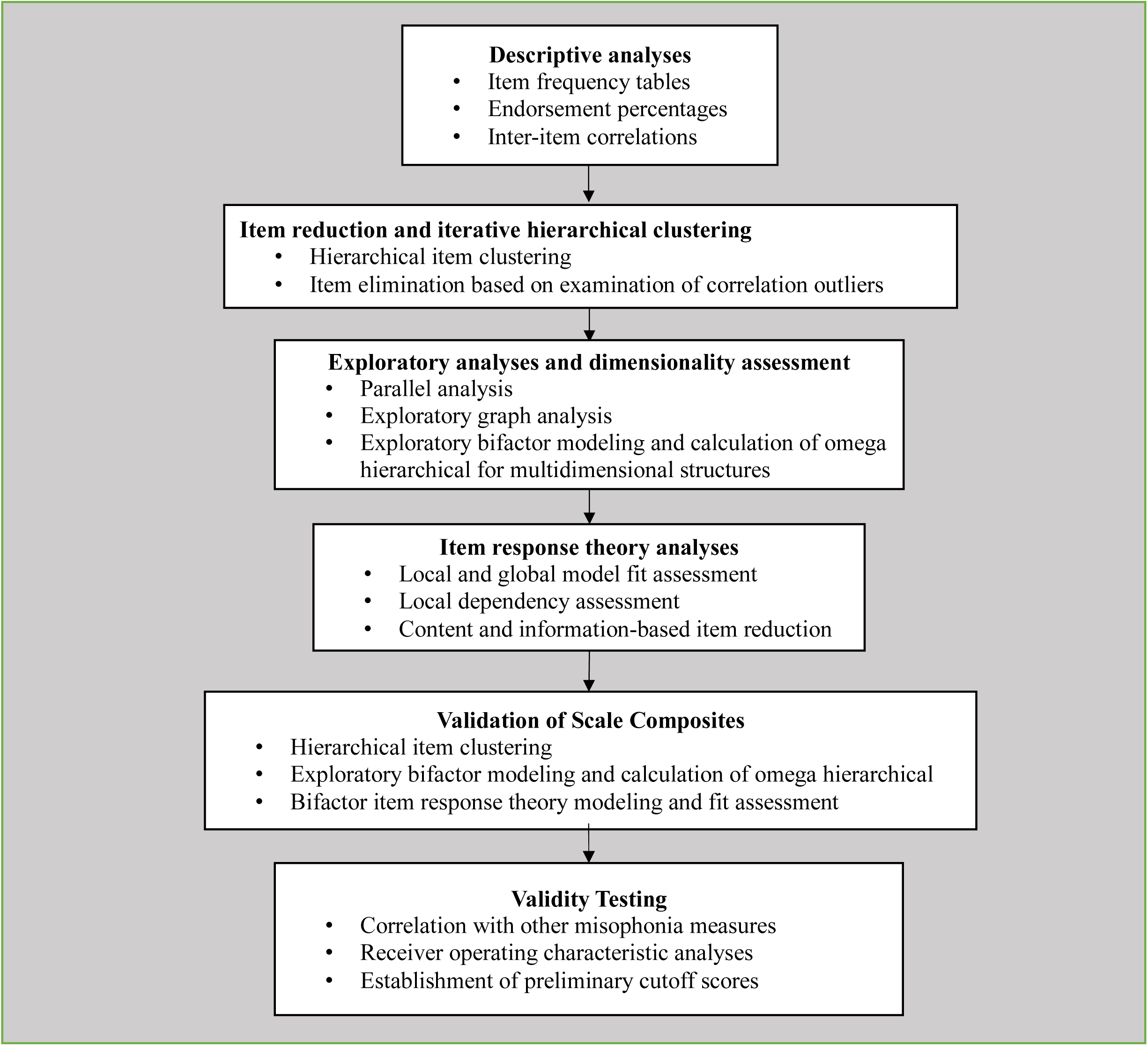
Scale refinement using Psychometric Analyses.

### Phase 1a: Development of Item Pool

#### Literature Review to Identify Existing Misophonia Measures

To begin identifying candidate items, a comprehensive literature search for measures explicitly purporting to measure misophonia was conducted. Measures identified included the Misophonia Questionnaire (MQ; Wu et al., 2014), Amsterdam Misophonia Scale (A-MISO-S; Schröder et al., 2013), Misophonia Assessment Questionnaire (MAQ; Johnson, 2014), Misophonia Activation Scale (MAS-1; Fitzmaurice, 2010), Misophonia Physiological Response Scale (MPRS; Bauman, 2015), Misophonia Coping Responses Scale (MCR; Johnson, 2014), Misophonia Trigger Severity Scale (MTS; Bauman; unpublished), and Misophonia Emotional Responses Scale (MER; Dozier, 2015). All items across these measures were reviewed and included in the initial pool. Next, redundant items were removed and several items were adapted to ensure consistency in tense (e.g., the item “*have violent thoughts”* from the MQ was replaced by “*I thought I want to be violent”*). These steps resulted in an initial pool of 75 candidate items.

#### Individual Interview

Based on the framework outlined above, a semi-structured interview was developed to ensure that the item pool would comprehensively capture the experience of individuals with misophonia. Two versions of the interview were created—one to be administered with individuals suffering from misophonia and the other with instructions adapted for family members of misophonia sufferers. Participants were included if they were 18 years or older and were reporting about the experience of a family member who was 13 years or older (when applicable). Additionally, criteria for “suffering from misophonia” (for participants themselves or their loved one/ family member), was a mean item score greater than 2 on the Misophonia Symptom Scale and Misophonia Emotions and Behaviors Scale and a score of 7 or higher on the Severity Scale of the Misophonia Questionnaire (Wu et al., 2014). This conservatively high level of misophonia symptoms was used in order to ensure that data were being collected from individuals characterized by a high level of symptom severity.

The interview contained a combination of close-and open-ended questions, assessing response to misophonic trigger sounds, ways of coping before, during and after the sound, impairment resulting from these symptoms and beliefs about the experience of misophonia. Close-ended questions, such as affective response to misophonic sounds, were designed using current psychometrically validated measures (e.g. PANAS; Mackinnon et al., 1999; Watson et al., 1988). Open-ended questions were developed by the authors based on clinical experience assessing and treating individuals with misophonia.

Participants were individuals with misophonia (*n* = 10) and family members of individuals with misophonia (*n* = 10). All participants were recruited via: (a) e-mail invitations to participants who had previously participated in our Center’s research and were interested in participating in future research, and (b) our Center’s website. Although there was no direct recruitment online made by study staff, it was determined through feedback from participants that the link to our Center website and link to participate was shared on various online and social media platforms.

Qualitative responses to each interview question were recorded verbatim by the interviewer using a standardized form. These data were then reviewed and for each question, and every unique response was recorded as a potential addition to the item pool. After identifying new items from interviews with sufferers and loved ones of those with misophonia, the resulting candidate item pool included 372 items.

### Phase 1b: Refinement of Initial Item Pool

#### Feedback on item wording, structure and applicability from misophonia sufferers

New participants (*n* = 10) were recruited who met the threshold for “clinical misophonia” on the MQ using the same recruitment and screening protocol as in Phase 1a. In addition to responding to the items themselves, participants provided free response feedback on each item and offered changes or additions to each subscale. Participants also gave feedback on the length and order of the scale, wording of instructions, and comprehensibility of items.

Each point of feedback was discussed by the study team. In response to this feedback, changes were made to enhance applicability and clarity of the general instructions and response options. Additionally, items flagged as confusing that were rarely endorsed (e.g., “I had psychic discomfort” and “I had sensory shock”) were removed.

#### Feedback from experts

The resulting item pool was sent to a group of professional experts. Expert feedback was reviewed by the study team and incorporated. Changes included clarification of instructions and inclusion of items considered relevant (e.g., added a question assessing types of sound triggers).

#### Study team qualitative review

Next, members of the study team reviewed all candidate items and made additional refinements to increase clarity and specificity of language. This included splitting of double-barreled items and further simplification of instructions and response options.

#### Analysis of literary demand

The reading level of each item and scale was assessed using both the Flesch-Kincaid Grade Level (FKGL) and Flesch Reading Ease (FRE) scores (Flesch, 1948; Kincaid et al., 1975), as implemented in the Readability Studio software package (Oleander Software, Ltd, Vandalia, OH, USA). To assure adequate readability by the majority of United States adults, we ensured that each subscale of the item pool had an average FKGL of 9 or below and an average FRE score of greater than 60 (i.e., “plain English”). Thus, when individual items or scales were above these benchmarks, changes were made to enhance readability for some items (e.g., “my muscles contracted automatically” changed to “I felt my muscles tense up or tighten”). After completion of all Phase 1b modifications, a final item pool of 377 items was prepared for Phase 2 data collection and psychometric analyses.

### Phase 2: Scale Refinement Using Psychometric Analyses

In Phase 2, the item pool was administered to a large sample in order to further refine and validate the measure using psychometric analyses.

#### Participants

Participants (*N* = 800) were recruited through Amazon’s Mechanical Turk (MTurk), a widely-used crowdsourcing platform that produces data of a similar or higher quality compared to other convenience samples (Hauser et al., 2019; Kees, Berry, Burton, & Sheehan, 2017; Miller et al., 2017). In accordance with MTurk best practices (Chandler & Shapiro, 2016), potential participants were unobtrusively screened for eligibility criteria of being between 18 to 65 years of age, English fluency and current residence in the U.S.

#### Data Integrity Check

In order to ensure data integrity, several stringent measures were taken. Participants were included only if they had a history on this platform of providing good quality responses (i.e., had an acceptance ratio > = 95%). Google’s reCAPTCHA was used to eliminate automated responses (i.e., “bots”). Additionally, a warning was included at the beginning of the survey to alert participants to the fact that their data quality would be monitored and compensation may not be given for poor-quality responses. Finally, three attention check questions were administered during the survey, and only participants who had accurate responses to all three were included in analyses. These measures are in line with recommendations for ensuring data quality for studies administered via MTurk (Chandler and Shapiro, 2016) and with measures taken by existing studies using this methodology (e.g., Everaert and Joorman, 2019). Indeed, several researchers have emphasized the need to eliminate careless responders, in order to reduce data quality problems including biased correlations among variables (Arias et al., 2020; Chandler et al., 2020). Of the 800 individuals initially recruited, a final sample of 424 participants (53%) passed the stringent data integrity checks and were included in the analyses.

#### Measures

##### Misophonia Questionnaire

(MQ; Wu et al., 2014). The MQ consists of 17 items that are divided into three subscales: the Misophonia Symptom Subscale, the Misophonia Emotions and Behaviors Subscale, and the Impairment Subscale. A cut score of seven (“Moderate”) or higher on the Impairment Subscale indicates clinically significant misophonia symptomology (Wu et al., 2014). Initial validation of the MQ demonstrated good internal consistency (*α* = .86 -.89; Wu al., 2014). Initial validation of the MQ demonstrated good internal consistency (*α* = .86 - .89; Wu

##### Amsterdam Misophonia Scale

(A-MISO-S; Schröder et al., 2013). The A-MISO-S is a semi-structured interview that probes six items: (1) the amount of time occupied by misophonia symptoms; (2) interference of misophonia in social functioning; (3) level of general distress (anger, disgust, irritation) the misophonia symptoms cause the individual; (4) how much effort it takes the individual to resist various misophonia impulses (i.e. thinking about the sound, diverting attention); (5) how much control the individual feels they have over their misophonia-related thoughts and anger; and (6) how much time an individual spends avoiding misophonia situations. In the current study, this measure was administered as a self-report questionnaire rather than a clinical interview. Scores from 0 to 4 are considered subclinical misophonic symptoms, 5–9 mild, 10–14 moderate, 15–19 severe, and 20–24 extreme. A good internal consistency of 0.81 was reported (Naylor et al., 2020). Reliability of the A-MISO-S total score in the current sample was good (*α* = .87).

##### Misophonia Activation Scale

(MAS-I; Fitzmaurice, 2010). The MAS-I is a two-item rating tool that focuses on the severity of emotional and physical responses to trigger sounds. No psychometric properties of the MAS-I have been published. Because it is only two items, reliability was not computed for the present study.

##### Misophonia Assessment Questionnaire

(MAQ; Johnson, 2014). The MAQ is a 21-item measure that assesses impact of misophonia. In one study (Dozier, 2015), the sum of the individual’s score in the MAQ was used to rate the severity of symptoms as mild (1-21), moderate (22-42), or severe (43-63). Psychometric properties of the MAQ have not been published. Reliability of the MAQ total score in the current sample was excellent (α = .96).

##### Misophonia Coping Responses Survey

(MCRS; Johnson, 2014). The MCRS is a 21-item scale that assesses how individuals cope with trigger sounds. No psychometric properties have been published. Reliability of the MCRS total score in the current sample was good (*α* = .89).

##### Misophonia Emotional Responses Scale

(MERS; Dozier, 2015). The MERS is a 26-item questionnaire of the frequency of emotional responses to misophonia triggers. Reliability of the MERS total score in the current sample was excellent (*α* = .94).

#### Data Analytic Plan

To refine the DMQ using psychometric analyses, we first split the item pool into nine subscales according to *a priori* theoretical item content groupings (“Affective Response,” “Physical Response,” “Cognitive Response,” “Behavioral Response,” “Coping–Before,” “Coping–During,” “Coping–After,” “Impairment,” and “Beliefs.” Each subscale was analyzed separately to enable item reduction and examination of item structure, using the following steps:

1. **Analysis of Response Rates and Item Redundancy.** Items for which greater than 90% of respondents marked “0” (never) or for which fewer than 2% of respondents marked either “3” or “4” (often or always/almost always) were removed. We also examined the matrix of inter-item (polychoric) correlations, flagging pairs of items with correlations greater than 0.8367 (i.e., sharing ≥70% of their variance). One item from each pair was removed, with items having more general content or higher endorsement rates retained for further analysis.
2. **Hierarchical Cluster Analysis.** The structure of remaining items within the subscale was explored using hierarchical cluster analysis of polychoric correlations (ICLUST; Revelle, 1979).

a. To reduce item redundancy, items that correlated so highly that they made artifactual “doublet” factors were flagged, and only a single item from the factor, with the higher endorsement rate or with greater theoretical meaning, was selected.
b. For larger clusters, inter-item correlations were calculated for all item pairs, and outlying correlations for each item were flagged using the MAD-Median rule (Wilcox & Rousselet, 2018). One item was then retained from each outlier pair based on degree of redundancy, percentage of endorsement, and theoretical meaning.
c. Items that resulted in a drop in Revelle’s (1979) beta coefficient ≥ 0.1 when added to a cluster were also removed, as the reduction in homogeneity implied that they were less relevant to the construct being measured. steps were repeated in several iterations after each set of items was removed, until no more items were flagged for removal.
3. **Exploratory Factor Analysis**. Parallel analysis (based on principal components and 500 column permutations as a null distribution; Lubbe, 2019) and exploratory graph analysis (Golino & Epskamp, 2017; Golino et al., 2020) were used to determine the appropriate number of factors. If these methods did not converge on a single number of factors, multiple solutions were examined and the most interpretable factor solution was selected. Exploratory factor analysis (EFA) was conducted for subscales with multidimensional structures using polychoric correlations, ordinary least squares extraction, and iterative target rotation techniques. An exploratory bifactor rotation (the iterative Schmid-Leiman procedure proposed by Garcia-Garzon et al., 2019) was performed on all subscales, and the omega hierarchical coefficient (ω_H_; Garcia-Garzon et al., 2020; Rodriguez et al., 2016; Revelle & Condon, 2019) was used to determine the proportion of scale variance attributable to a general factor. Values of ω_H_ greater than 0.8 (indicating >80% of scale variance attributable to the general factor) were taken to support the appropriateness of a bifactor structure (Rodriguez et al., 2016). In cases where a one-factor solution was optimal, the exploratory analysis was omitted, and we proceeded directly to item response theory modeling. Items deemed peripherally related to the constructs of interest based on EFA results (e.g., several items that form a specific factor but have relatively low general factor loadings) were removed at this stage.
4. **Item Response Theory Analysis.** Item Response Theory (IRT) was used to evaluate global model fit and eliminate local dependence or ill-fitting items. We fit a graded response model (Samejima, 1969) to each unidimensional subscale and a bifactor graded response model (Cai et al., 2011) to subscales with a bifactor structure using maximum marginal likelihood estimation via the Bock–Aitkin EM algorithm (Bock & Aitkin, 1981), as implemented in the *mirt* R package (Chalmers, 2012). The overall fit of the model was tested using the *C*_2_ statistic (Cai & Monro, 2014), *C*_2_-based Comparative Fit Index (CFI_C2_), *C*_2_-based Root Mean Square Error of Approximation (RMSEA_C2_), and the Standardized Root Mean Square Residual (SRMR). Although no absolute fit index cutoffs were used, the guidelines provided by Maydeu-Olivares & Joe (2014) were used to qualitatively interpret IRT model fit indices. The assumption of local item independence was tested using the standardized local dependence chi-squared statistic (LD-χ^2^; Chen & Thissen, 1997), with χ^2^ 10 indicating significant local dependence (Toland et al., 2017). To assess local model misfit, we examined residual correlations for each model, with values greater than ±0.1 indicative of significant local strain. In cases where an item pair was flagged for significant local dependence or local item misfit, we removed one of the offending items, retaining the item with more general content or fewer large residual correlations. After item removal, the IRT model was re-fit and examined using the same indices until a model showed good overall fit, no local dependence, and minimal local strain.
5. **Item Reduction and Scale Refinement.** The resulting item set fit by the IRT model was further refined to reduce the overall number of items while preserving broad content coverage. In accordance with recommendations by Goetz et al. (2013), we utilized a combination of statistical analyses and judgments based on item content to determine the items to retain. Examining the item information curves derived from each IRT model, we chose a reduced set of 5–15 items to form each final subscale. In particular, we prioritized items with maximal information in the range θ = [0, 3], as we felt that discrimination between individuals in this range of latent misophonia trait levels would be most informative for clinical and research use. However, we did not simply retain the items with the highest factor loadings or information values, as doing so often serves to increase scale homogeneity at the cost of content validity (Clifton, 2020). Instead, we selected items with high information compared to other items with similar content in order to balance high levels of scale information with low item content overlap (Edelen & Reeve, 2007). Once a theoretically justifiable reduced set of items was selected, this item set was fit to a unidimensional or bifactor graded response model, and fit was examined using *C*_2_, CFI_C2_, RMSEA_C2_, SRMR, and examination of residual correlations. The final model for each reduced subscale was required to exhibit adequate global fit (i.e., approximately CFI_C2_ > 0.95, RMSEA_C2_ < 0.089, SRMR < 0.05), no local dependency, and adequate representation of all content areas included in the item pool first submitted to IRT analyses. When the subscale was deemed by the study team to have insufficient item content in specific areas, one or more additional items that had been removed from the pool due to high inter-item correlations were provisionally re-introduced to the pool of potential scale items, and analyses were repeated iteratively until the resulting set of items met our criteria of adequate measurement model fit and sufficient content breadth.

### Inter-scale Correlations

Pearson correlations between the various subscales were analyzed in order to describe the relationships between the assessed constructs and further justify the conceptual groupings of certain scales into multidimensional composites.

### Composite Scale Analyses

Once item sets were developed for all subscales, we further analyzed the utility of two theoretically meaningful multidimensional composite scales: The Duke Misophonia Symptoms Scale (comprised of the “Affective Response,” “Physical Response,” and “Cognitive Response” subscales) and the Duke Misophonia Coping Scale (comprised of the three “Coping” subscales representing strategies before, during, and after exposure to a trigger sound). To analyze the hierarchical structure of each composite scale, we used the ICLUST algorithm (on polychoric correlations) to examine the ways in which items formed sub-clusters before combining into the overall scale composite. We then conducted an EFA, using a direct full-rank bifactor rotation (BiFAD; Giordano & Waller, 2019; Waller, 2018) with a pre-specified target matrix of 0s and 1s defined by the sub-clusters of the ICLUST solution. The BiFAD solution was then used as input to the iterative Schmid–Leiman algorithm (Garcia-Garzon et al., 2019), which converged on the final EFA model for each composite. Coefficient ω_H_ was again used to estimate the proportion of general factor variance, with values greater than 0.8 supporting the validity of a total score for each composite scale. The EFA loadings matrices was then used to construct a bifactor IRT model, with items loading on all factors on which their EFA loadings exceeded 0.2. The IRT model for each composite scale was a bifactor graded response model, fit using the Metropolis-Hastings-Robbins-Monro algorithm due to computational limitations of the EM algorithm with high-dimensional models (Cai et al., 2011). Final IRT models for composite scales were required to have a lack of local item dependency and severe local strain (i.e., any residual correlations greater than ±0.15). Fit was further quantified using *C*_2_, CFI_C2_, RMSEA_C2_, and SRMR, with the guidelines provided by Maydeu-Olivares & Joe (2014) used to qualitatively interpret these indices.

### External Validation Using Existing Measures

External validation (i.e., degree to which the relations between the instrument and external criteria are consistent with theory) was examined by estimating correlations with external constructs. This involved the following steps:

1. Correlations between the DMQ and existing measures of misophonia were examined as evidence of convergent validity.
2. Composite and Scale scores of the DMQ were examined to test their ability to discriminate between individuals with “clinical” or “non-clinical” misophonia, based on criteria determined by the Misophonia Questionnaire (i.e., score of ≥ 7 on the severity subscale, Wu et al., 2014). This was chosen in light of the existing literature using this criterion to assess moderate or higher misophonia severity.

## Results

Participants were 54.5% male, and predominantly White (see Table 1). The mean item score on the MQ Symptom Scale was 1.18 (SD = 0.78) and on the MQ Emotions and Behavior Scale was 1.08 (SD = 0.64), suggesting that on average participants marked responses between “rarely true” and “sometimes true” when asked how often they were sensitive to various potential sound triggers, and how often they experienced certain emotional and behavioral responses to these triggers. Responses to the MQ Impairment Scale (*M* = 4.21, *SD* = 2.52) suggest that on average the sample reported “mild sound sensitivities.” Sixty-seven participants (15.8%) reported “clinically significant symptoms” on the MQ (i.e., ≥ 7 on the impairment subscale (Wu et al. 2014).

**Table 1.**
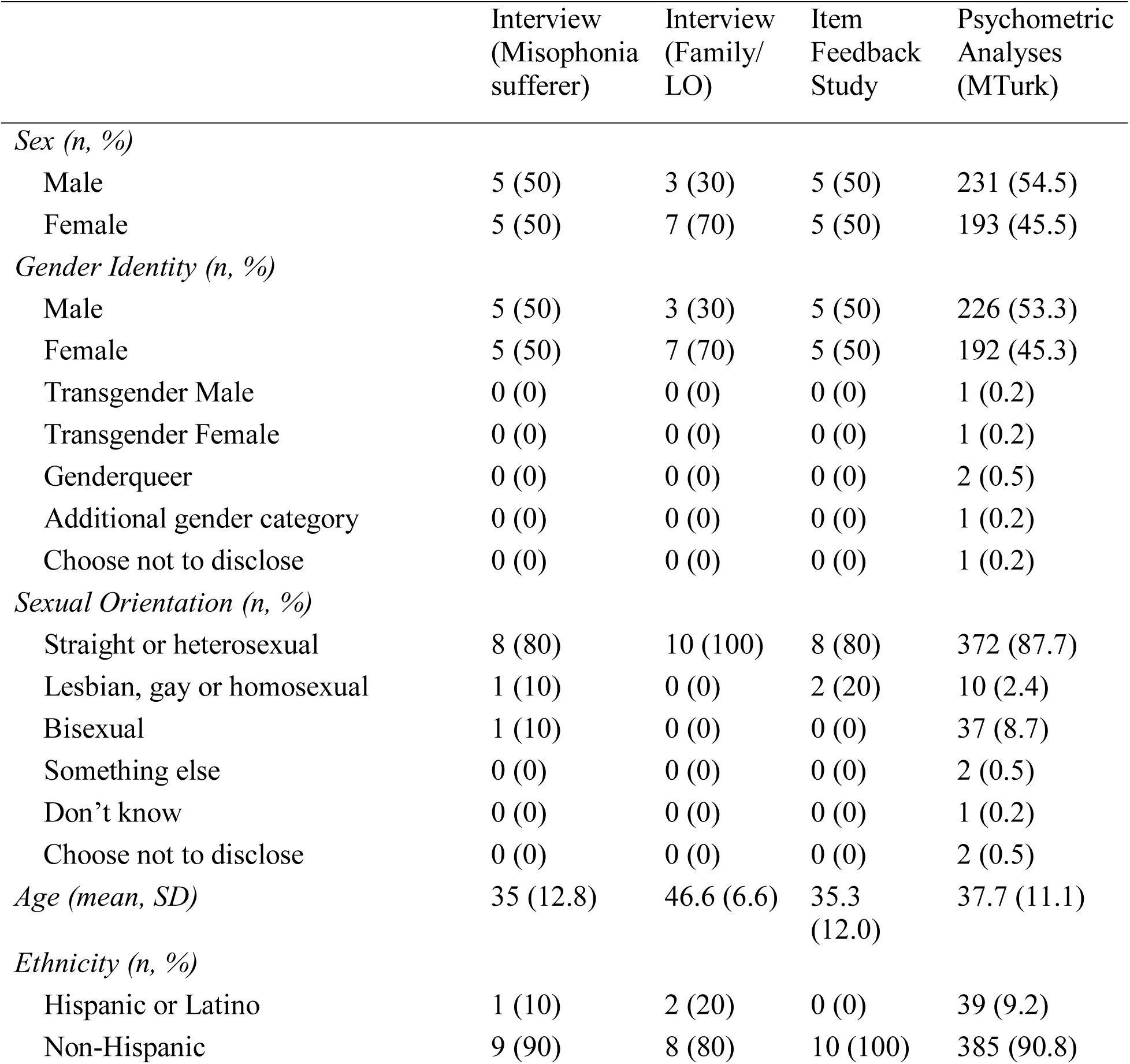
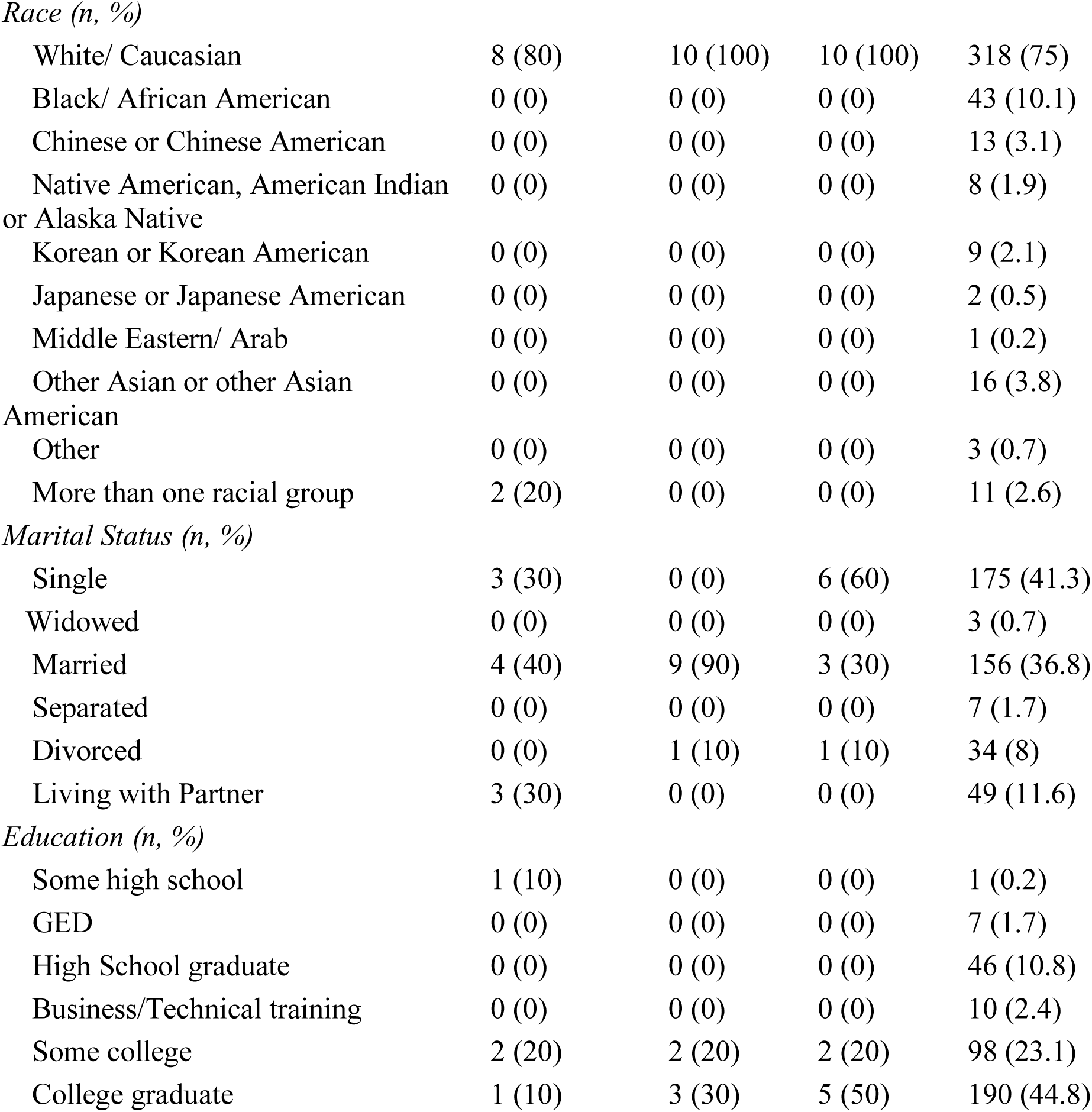
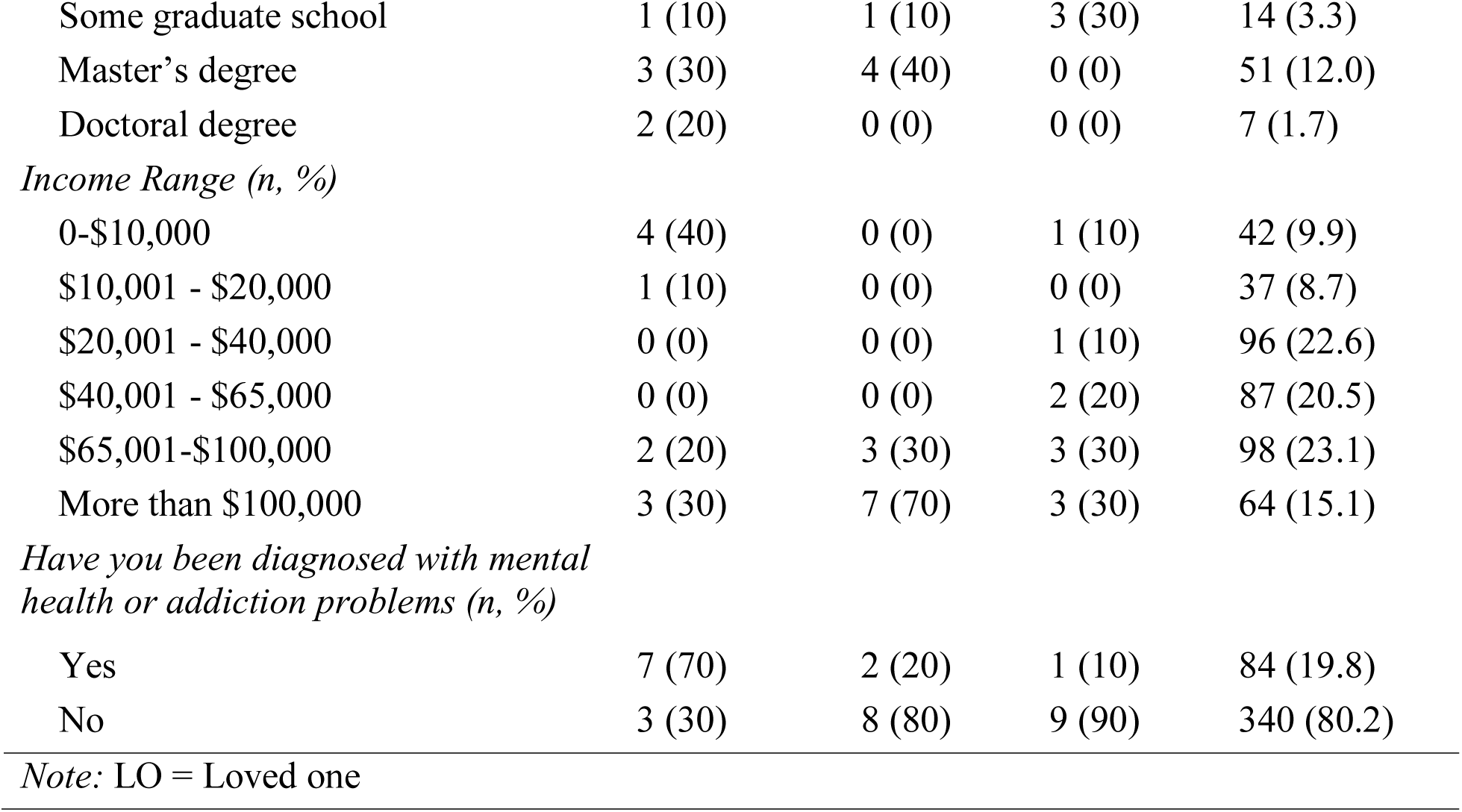
Demographic Data.

|  | Interview<br>(Misophonia<br>sufferer) | Interview<br>(Family/<br>LO) | Item<br>Feedback<br>Study | Psychometric<br>Analyses<br>(MTurk) |
| --- | --- | --- | --- | --- |
| *Sex (n, %)* |  |  |  |  |
| Male | 5 (50) | 3 (30) | 5 (50) | 231 (54.5) |
| Female | 5 (50) | 7 (70) | 5 (50) | 193 (45.5) |
| *Gender Identity (n, %)* |  |  |  |  |
| Male | 5 (50) | 3 (30) | 5 (50) | 226 (53.3) |
| Female | 5 (50) | 7 (70) | 5 (50) | 192 (45.3) |
| Transgender Male | 0 (0) | 0 (0) | 0 (0) | 1 (0.2) |
| Transgender Female | 0 (0) | 0 (0) | 0 (0) | 1 (0.2) |
| Genderqueer | 0 (0) | 0 (0) | 0 (0) | 2 (0.5) |
| Additional gender category | 0 (0) | 0 (0) | 0 (0) | 1 (0.2) |
| Choose not to disclose | 0 (0) | 0 (0) | 0 (0) | 1 (0.2) |
| *Sexual Orientation (n, %)* |  |  |  |  |
| Straight or heterosexual | 8 (80) | 10 (100) | 8 (80) | 372 (87.7) |
| Lesbian, gay or homosexual | 1 (10) | 0 (0) | 2 (20) | 10 (2.4) |
| Bisexual | 1 (10) | 0 (0) | 0 (0) | 37 (8.7) |
| Something else | 0 (0) | 0 (0) | 0 (0) | 2 (0.5) |
| Don't know | 0 (0) | 0 (0) | 0 (0) | 1 (0.2) |
| Choose not to disclose | 0 (0) | 0 (0) | 0 (0) | 2 (0.5) |
| *Age (mean, SD)* | 35 (12.8) | 46.6 (6.6) | 35.3<br>(12.0) | 37.7 (11.1) |
| *Ethnicity (n, %)* |  |  |  |  |
| Hispanic or Latino | 1 (10) | 2 (20) | 0 (0) | 39 (9.2) |
| Non-Hispanic | 9 (90) | 8 (80) | 10 (100) | 385 (90.8) |
| *Race (n, %)* |  |  |  |  |
| White/ Caucasian | 8 (80) | 10 (100) | 10 (100) | 318 (75) |
| Black/ African American | 0 (0) | 0 (0) | 0 (0) | 43 (10.1) |
| Chinese or Chinese American | 0 (0) | 0 (0) | 0 (0) | 13 (3.1) |
| Native American, American Indian<br>or Alaska Native | 0 (0) | 0 (0) | 0 (0) | 8 (1.9) |
| Korean or Korean American | 0 (0) | 0 (0) | 0 (0) | 9 (2.1) |
| Japanese or Japanese American | 0 (0) | 0 (0) | 0 (0) | 2 (0.5) |
| Middle Eastern/ Arab | 0 (0) | 0 (0) | 0 (0) | 1 (0.2) |
| Other Asian or other Asian<br>American | 0 (0) | 0 (0) | 0 (0) | 16 (3.8) |
| Other | 0 (0) | 0 (0) | 0 (0) | 3 (0.7) |
| More than one racial group | 2 (20) | 0 (0) | 0 (0) | 11 (2.6) |
| *Marital Status (n, %)* |  |  |  |  |
| Single | 3 (30) | 0 (0) | 6 (60) | 175 (41.3) |
| Widowed | 0 (0) | 0 (0) | 0 (0) | 3 (0.7) |
| Married | 4 (40) | 9 (90) | 3 (30) | 156 (36.8) |
| Separated | 0 (0) | 0 (0) | 0 (0) | 7 (1.7) |
| Divorced | 0 (0) | 1 (10) | 1 (10) | 34 (8) |
| Living with Partner | 3 (30) | 0 (0) | 0 (0) | 49 (11.6) |
| *Education (n, %)* |  |  |  |  |
| Some high school | 1 (10) | 0 (0) | 0 (0) | 1 (0.2) |
| GED | 0 (0) | 0 (0) | 0 (0) | 7 (1.7) |
| High School graduate | 0 (0) | 0 (0) | 0 (0) | 46 (10.8) |
| Business/Technical training | 0 (0) | 0 (0) | 0 (0) | 10 (2.4) |
| Some college | 2 (20) | 2 (20) | 2 (20) | 98 (23.1) |
| College graduate | 1 (10) | 3 (30) | 5 (50) | 190 (44.8) |
| Some graduate school | 1 (10) | 1 (10) | 3 (30) | 14 (3.3) |
| Master's degree | 3 (30) | 4 (40) | 0 (0) | 51 (12.0) |
| Doctoral degree | 2 (20) | 0 (0) | 0 (0) | 7 (1.7) |
| *Income Range (n, %)* |  |  |  |  |
| 0-\$10,000 | 4 (40) | 0 (0) | 1 (10) | 42 (9.9) |
| \$10,001 - \$20,000 | 1 (10) | 0 (0) | 0 (0) | 37 (8.7) |
| \$20,001 - \$40,000 | 0 (0) | 0 (0) | 1 (10) | 96 (22.6) |
| \$40,001 - \$65,000 | 0 (0) | 0 (0) | 2 (20) | 87 (20.5) |
| \$65,001-\$100,000 | 2 (20) | 3 (30) | 3 (30) | 98 (23.1) |
| More than \$100,000 | 3 (30) | 7 (70) | 3 (30) | 64 (15.1) |
| *Have you been diagnosed with mental health or addiction problems (n, %)* |  |  |  |  |
| Yes | 7 (70) | 2 (20) | 1 (10) | 84 (19.8) |
| No | 3 (30) | 8 (80) | 9 (90) | 340 (80.2) |

*Note:* LO = Loved one

For participants (*n* = 35) who responded “I am not bothered by any sounds or sights at are as associated with sounds,” “Not Applicable” responses to following questions were transformed into “0”. Further, total scores for each subscale were computed for all participants, with items marked “Not Applicable” imputed as the mean of all other items on the subscale. These changes were made to help interpret the “Not Applicable” responses in subsequent analyses.

### Scale Specific Item Reduction

#### Affective Response Subscale

The initial affective response scale contained 31 items (See Supplemental Table), eight of which were removed due to low item endorsement. The remaining 23 items were subjected to three rounds of hierarchical clustering and item removal, during which an additional six items were removed. The remaining 17 items formed three clusters, which were interpreted as “Anger/Disgust” (10 items), “Anxiety” (four items) and “Sadness/Disengagement” (three items). EGA supported the three-cluster solution by suggesting three latent dimensions, but parallel analysis suggested that only two be retained. Thus, bifactor solutions with two and three specific factors were examined. The three-factor solution produced a loadings matrix that approximated the three clusters identified by ICLUST, and thus this solution was retained. General factor saturation was strong (ω_H_ = 0.909), indicating that a bifactor structure was appropriate for this subscale. However, based on examination of the factor loadings and percentage of item variance accounted for by the general factor, the “Sadness/Disengagement” cluster of items seemed peripherally related to the remaining items and were omitted from the IRT analyses.

The remaining 14 items were then fit to a bifactor graded response model with two specific factors for the 10 “Anger/Disgust” items and four “Anxiety” items, respectively. The model showed acceptable fit (*C*_2_(63) = 142.5, *p* < 0.001, CFI_C2_ = 0.992, RMSEA_C2_ = 0.055, SRMR = 0.036), no local dependence, and only a single large residual. Using general factor item information curves and content judgments, we removed six additional items, settling on an eight-item affect scale (five Anger/Disgust items, three Anxiety items). A bifactor graded response model exhibited adequate fit to data from these items (*C*_2_(12) = 35.3, *p* < 0.001, CFI_C2_ =0.992, RMSEA_C2_ = 0.068, SRMR = 0.040), as well as no local dependence (all LD-χ^2^ < 3.55) and no residuals indicative of local strain. Marginal reliability for the general factor score was strong (ρ_xx_ = 0.884).

#### Physical Response Subscale

The initial physical response scale contained 33 items (See Supplemental Table), 13 of which were removed due to low item endorsement. The remaining 20 items were subjected to three rounds of hierarchical clustering and item removal, during which an additional five items were removed. The remaining 15 items formed a single cluster, and both parallel analysis and EGA suggested a single factor was sufficient to describe these data. We thus proceeded directly to IRT analysis.

A unidimensional graded response model was fit to the 15 remaining physical response items. Model fit was borderline acceptable (*C*_2_(90) = 334.6, *p* < 0.001, CFI_C2_ = 0.971, RMSEA_C2_ = 0.081, SRMR = 0.061), and while no local dependence was present, 10 residuals exceed the cutoff of ±0.1. Three items were removed, and the unidimensional model with the remaining 12 items exhibited improved fit (*C*_2_(54) = 154.8, *p* < 0.001, CFI_C2_ = 0.983, RMSEA_C2_ = 0.067, SRMR = 0.047), no local dependence (all LD-χ^2^ < 3.63), and no residuals indicative of local strain. A reduced set of five items was derived from this 12-item subscale using a combination of content analysis and item information curves. The resulting scale exhibited excellent fit to a unidimensional model (*C*_2_(5) = 7.84, *p* = 0.165, CFI_C2_ = 0.997, RMSEA_C2_ = 0.037, SRMR = 0.026), no local dependence (all LD-χ^2^ < 1.56), and no large residuals. Marginal reliability for the scale’s factor score was acceptable (ρ_xx_ = 0.739).

#### Cognitive Response Subscale

The initial cognitive response scale contained 67 items (Supplemental Table), seven of which were removed due to extremely high inter-item correlations (*r* > 0.837). The remaining 60 items were subjected to eight rounds of hierarchical clustering and item removal, during which an additional 29 items were removed. The remaining 31 items formed four clusters, which were interpreted as “Aggression” (three items), “Self-criticism” (five items), “Despair” (11 items), and “Escape” (12 items). EGA supported the four-cluster solution by suggesting four latent dimensions, but parallel analysis suggested that only three be retained. Thus, bifactor solutions with three and four specific factors were examined. The four-factor solution produced a loadings matrix that approximated the clusters identified by ICLUST, and thus this solution was retained. General factor saturation was strong (ω_H_ = 0.858), supporting the bifactor structure.

The 31-item cognitive response scale was fit to a bifactor graded response model, with four factors derived from EFA loadings. The global fit of this model was acceptable (*C*_2_(401) = 746.8, *p* < 0.001, CFI_C2_ = 0.991, RMSEA_C2_ = 0.046, SRMR = 0.046), and no local dependence was present. However, 15 residual correlations exceeded ±0.1, indicating substantial amounts of local misfit. Examination of these correlations suggested that the model was substantially over-estimating the correlations between “Self-criticism” items and “Aggression” items, indicating that these two domains should not load onto the same general factor. Given the theoretical importance of aggression to the conceptualization of misophonia (e.g., Jager et al., 2020) and the presence of self-critical content on the DMQ “Beliefs” scale, we chose to eliminate the “Self-criticism” from the cognitive response scale, retaining only items from the other three specific factors. The item pool was further reduced based on item information curves and content balance, resulting in a 10-item scale with items in the three domains of “Aggression” (three items), “Despair” (four items), and “Escape” (three items). A bifactor model with those three specific factors fit the data extremely well (*C*_2_(23) = 30.1, *p* = 0.148, CFI_C2_ = 0.998, RMSEA_C2_ = 0.027, SRMR = 0.030), and the resulting scale had no local dependence (all LD-χ^2^ < 2.72) or large residual correlations. Marginal reliability for the general factor score was good (ρ_xx_ = 0.894).

#### Behavioral Response Subscale

The initial behavioral response scale contained 43 items (See Supplemental Table), 10 of which were removed due to low item endorsement. The remaining 33 items were subjected to five rounds of hierarchical clustering and item removal, during which an additional 19 items were removed. The remaining 14 items formed a single cluster, and both parallel analysis and EGA suggested a single factor was sufficient to describe these data. We thus proceeded directly to IRT analysis.

A unidimensional graded response model was fit to the 14 remaining behavioral response items. Model fit was borderline acceptable (*C*_2_(54) = 162.0, *p* < 0.001, CFI_C2_ = 0.967, RMSEA_C2_ = 0.071, SRMR = 0.068), with one locally dependent item pair and seven residuals exceeding ±0.1. Iterative item refinement based on content balance, item information curves, and fit of the resulting model caused an additional nine items to be removed. The unidimensional model with the remaining five items exhibited much improved fit (*C*_2_(5) = 11.7, *p* = 0.040, CFI_C2_ = 0.989, RMSEA_C2_ = 0.057, SRMR = 0.040), no local dependence (all LD-χ^2^ < 2.54), and no residuals indicative of local strain. Marginal reliability for the scale’s factor score was acceptable (ρ_xx_ = 0.748).

#### Coping-before Subscale

The initial coping-before scale contained 23 items (See Supplemental Table), none of which were removed due to low item endorsement or high initial correlations. These 23 items were subjected to four rounds of hierarchical clustering and item removal, during which an additional nine items were removed. The remaining 14 items formed a single cluster, and both parallel analysis and EGA suggested a single factor was sufficient to describe these data. We thus proceeded directly to IRT analysis.

A unidimensional graded response model was fit to the 14 remaining coping-before items. Model fit was borderline acceptable (*C*_2_(77) = 258.5, *p* < 0.001, CFI_C2_ = 0.971, RMSEA_C2_ = 0.076, SRMR = 0.073), with one locally dependent item pair and 18 residuals exceeding ±0.1. Iterative item refinement based on content balance, item information curves, and fit of the resulting model caused an additional eight items to be removed. The unidimensional model with the remaining six items displayed excellent fit (*C*_2_(9) = 14.8, *p* = 0.095, CFI_C2_ = 0.996, RMSEA_C2_ = 0.040, SRMR = 0.037), no local dependence (all LD-χ^2^ < 5.94), and no residuals indicative of local strain. Marginal reliability for the scale’s factor score was good (ρ_xx_ = 0.825).

#### Coping-during Subscale

The initial coping-during scale contained 33 items (See Supplemental Table), five of which were removed due to low item endorsement (*n*=4) or very high inter-item correlations (*n*=1). The remaining 28 items were subjected to four rounds of hierarchical clustering and item removal, during which an additional 15 items were removed. The remaining 13 items formed a single cluster with three defined sub-clusters, which were interpreted as “Cognitive Reframing” (four items), “Masking” (four items), and “Distraction/Active Coping” (five items). EGA supported the three-cluster solution by suggesting three latent dimensions, but parallel analysis suggested that only a single factor be retained. Thus, we fit a bifactor solution with three specific factors to the data. General factor saturation was strong (ω_H_ = 0.832), supporting the bifactor structure.

The 13-item coping-during scale was fit to a bifactor graded response model, with three specific factors derived from EFA loadings. The global fit of this model was borderline acceptable (*C*_2_(52) = 100.7, *p* < 0.001, CFI_C2_ = 0.989, RMSEA_C2_ = 0.048, SRMR = 0.057), and no local dependence was present. Four residual correlations exceeded ±0.1, indicating a small amount of local misfit. Iterative item refinement based on content balance, item information curves, and fit of the resulting model cause an additional three items to be removed. This resulted in a 10-item scale with items in the three domains of “Masking” (four items), “Distraction” (three items), and “Cognitive Techniques” (three items). A bifactor model with those three specific factors displayed good fit to the data (*C*_2_(25) = 50.7, *p* = 0.002, CFI_C2_ = 0.991, RMSEA_C2_ = 0.050, SRMR = 0.048). Additionally, the resulting scale exhibited no local dependence (all LD-χ^2^ < 6.70), and one single residual correlation greater than ±0.1. Marginal reliability for the general factor score was good (ρ_xx_ = 0.877).

#### Coping-after Subscale

The initial cognitive response scale contained 25 items (See Supplemental Table), three of which were removed due to low item endorsement (*n*=2) or very high inter-item correlations (*n*=1). The remaining 22 items were subjected to five rounds of hierarchical clustering and item removal, during which an additional 14 items were removed. The remaining eight items formed a single cluster, and both parallel analysis and EGA suggested a single factor was sufficient to describe these data. We thus proceeded directly to IRT analysis.

A unidimensional graded response model was fit to the eight remaining coping-after items. Model fit was borderline acceptable (*C*_2_(20) = 63.9, *p* < 0.001, CFI_C2_ = 0.979, RMSEA_C2_ = 0.073, SRMR = 0.059), with no local dependence and four residuals exceeding ±0.1. Iterative item refinement based on content balance, item information curves, and fit of the resulting model cause an additional three items to be removed. The unidimensional model with the remaining five items displayed excellent fit (*C*_2_(5) = 9.24, *p* = 0.100, CFI_C2_ = 0.996, RMSEA_C2_ = 0.045, SRMR = 0.046), no local dependence (all LD-χ^2^ < 5.67), and no residuals indicative of local strain. Marginal reliability for the scale’s factor score was acceptable (ρ_xx_ = 0.783).

#### Impairment Subscale

The initial impairment scale contained 43 items (See Supplemental Table), 19 of which were removed due to low item endorsement (*n*=2) or very high inter-item correlations (*n*=17). The remaining 24 items were subjected to three rounds of hierarchical clustering and item removal, during which an additional 10 items were removed. The remaining 14 items formed a single cluster with three defined sub-clusters, which were interpreted as “Relationship Problems” (five items), “Self-confidence” (four items), and “Occupational Problems” (four items). EGA supported the three-cluster solution by suggesting four latent dimensions (the fourth dimension consisting of two items that fit poorly into the other three subclusters), but parallel analysis suggested that only a single factor be retained. Thus, we fit a bifactor solution with three specific factors to the data. General factor saturation was very strong (ω_H_ = 0.915), supporting the bifactor structure. Additionally, the “Self-confidence” factor was essentially subsumed into the general factor, leaving only the specific occupational and relationship factors as contributing meaningful amounts of additional variance. Thus, when constructing a confirmatory IRT model for this scale, we only included specific factors for relationship and occupational problems, with the remainder of the items loading on the general factor alone.

The 14-item impairment scale was fit to a bifactor graded response model, with five items loading solely on the general factor, five items loading on a specific “Relationship Problems” factor, and four items loading on a specific “Occupational Problems” factor. The global fit of this model was acceptable (*C*_2_(52) = 100.7, *p* < 0.001, CFI_C2_ = 0.989, RMSEA_C2_ = 0.048, SRMR = 0.049), and no local dependence was present. Four residual correlations exceeded ±0.1, indicating a small amount of local misfit. Iterative item refinement based on content balance, item information curves, and fit of the resulting model cause an additional two items to be removed. This resulted in a 12-item scale with items in the domains of “Relationship Problems” (five items), “Occupational Problems” (four items), and “Other Impairment” (general factor loadings only; three items). A bifactor model with specific factors for relationship and occupational problems displayed good fit to the data (*C*_2_(45) = 75.37, *p* = 0.003, CFI_C2_ = 0.996, RMSEA_C2_ = 0.042, SRMR = 0.040). Additionally, the resulting scale exhibited no local dependence (all LD-χ^2^ < 2.39), and no residual correlations greater than ±0.1. Marginal reliability for the general factor score was good (ρ_xx_ = 0.800).

### Beliefs Subscale

The initial beliefs scale contained 43 items (See Supplemental Table), nine of which were removed due very high inter-item correlations. The remaining 34 items were subjected to seven rounds of hierarchical clustering and item removal, during which an additional 18 items were removed. The remaining 16 items formed two clusters with three defined sub-clusters, which were interpreted as “Self-criticism” (six items), “Self-pity” (10 items). EGA and parallel analysis both suggested two latent dimensions, further supporting the two-cluster solution. Thus, we fit a bifactor solution with two specific factors to the data. General factor saturation was strong (ω_H_ = 0.821), providing additional support for the bifactor structure.

The 16-item beliefs scale was fit to a bifactor graded response model, with two specific factors derived from EFA loadings. The global fit of this model was borderline acceptable (*C*_2_(88) = 213.1, *p* < 0.001, CFI_C2_ = 0.990, RMSEA_C2_ = 0.060, SRMR = 0.057). However, one item pair was locally dependent and 10 residual correlations exceeded ±0.1, indicating substantial local misfit. Iterative item refinement based on content balance, item information curves, and fit of the resulting model cause an additional six items to be removed and four items that were previously removed from the item pool to be added back in. This resulted in a 14-item scale, which was modeled as a bifactor model with four “Self-criticism” items loading onto a single group factor and the remaining 10 items loading solely onto the general factor (i.e., a bifactor S–1 model; Burns et al., 2020; Eid et al., 2017). This bifactor S–1 displayed good fit to the data (*C*_2_(73) = 113.0, *p* = 0.002, CFI_C2_ = 0.996, RMSEA_C2_ = 0.037, SRMR = 0.041). Additionally, the resulting scale exhibited no local dependence (all LD-χ^2^ < 5.00), and a single residual correlation greater than ±0.1. Marginal reliability for the general factor score was good (ρ_xx_ = 0.882).

### Refinement of Item Responses for “Sound Type” and “Frequency” Questions

Two questions – one assessing the *type* of sound that is bothersome, and another assessing the frequency with which a person was bothered by a sound, were administered at the beginning of the measure.

*Sound type:* All the response options were endorsed, with an endorsement rate of higher than 5% (Table 2), therefore, no option was eliminated on the basis of low endorsement rate. When examining the correlations among the response options, three pairs of items were identified as potentially redundant due to inter-item tetrachoric correlations greater than 0.5 [*People making nasal sounds* and *People making throat sounds* (*r* = 0.579); *Animal making repetitive sounds* and *Animals making eating sounds* (*r* = 0.550); and *Seeing a sound produced without being able to hear it* and *Seeing someone do anything that might make a bothersome sound, before the sound occurs* (*r* = 0.512)]. Each of these item pairs was thus collapsed into a single item encompassing content from both items [e.g., nasal and throat sounds items were combined into *People making nasal/throat sounds (e.g., sniffing, sneezing, nose-whistling, coughing, throat-clearing)*].

**Table 2.**
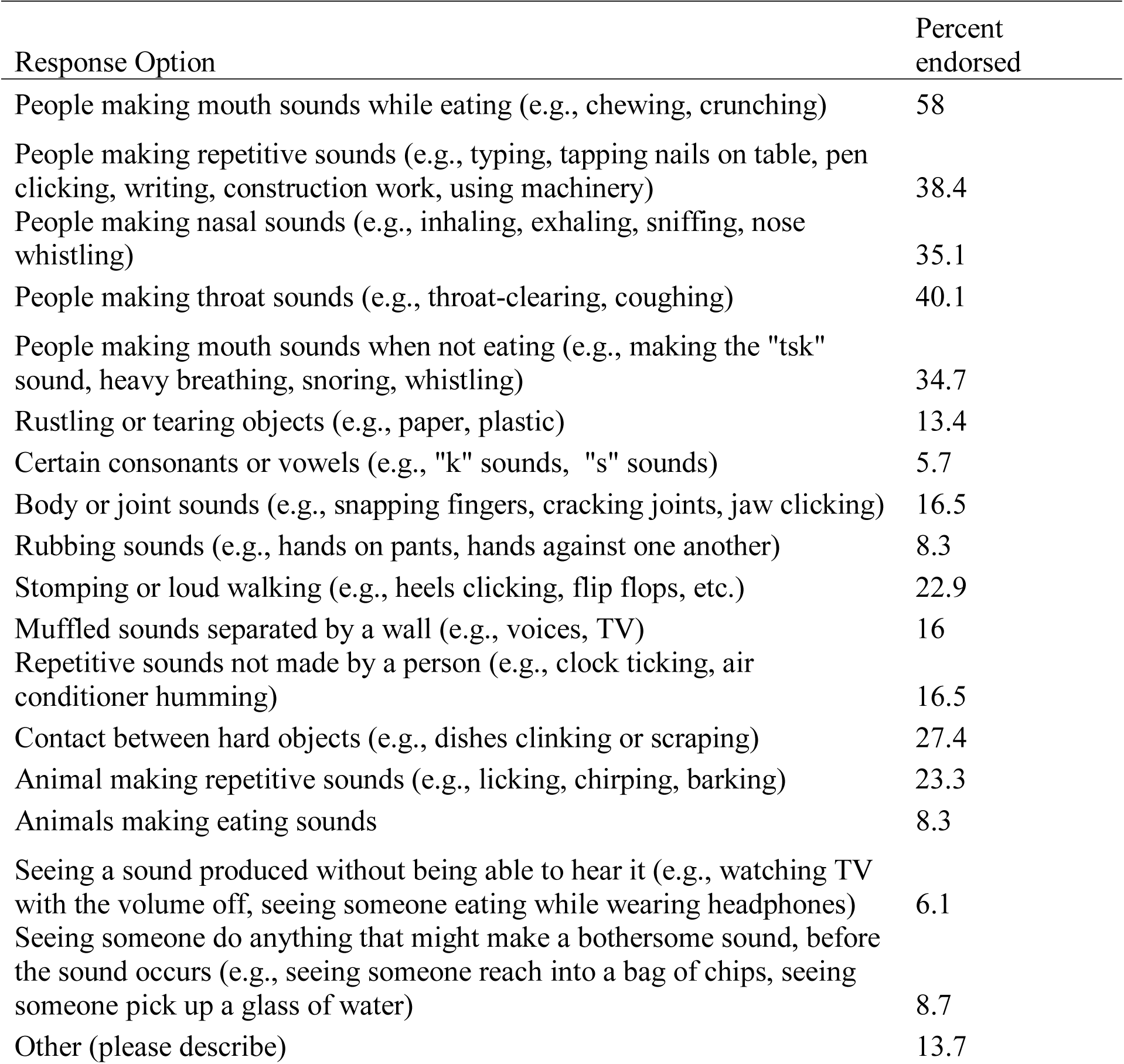
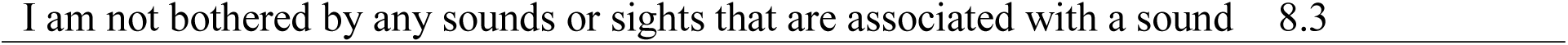
Endorsement Rates for Duke Misophonia Questionnaire Sound Type by Item.

Participants were allowed to type in sounds not captured by the listed response options, under “other”. These qualitative responses were analyzed and separated into four categories based on the authors’ assessment– (a) responses that could be included under existing options, without modifying the category, (b) responses that could be included in existing options by broadening the options, (c) responses that were considered very specific, and therefore best captured under the *other* response, and (d) responses that implied distress about the loudness of the sound, more so than the sound itself, which could conflate the experience with hyperacusis (Fackrell et al., 2019). Based on this categorization, some existing response options were broadened (e.g. *Speech sounds* was added as an option instead of *Certain consonants or vowels, Styrofoam rubbing together* was added as an example of *Rubbing sounds*).

Finally, given that several participants included sounds under “other” that may be distressing to *most* individuals, regardless of whether they have misophonia (e.g. “baby crying”, “fingernails on chalkboard”) the authors decided to modify the instructions to include comparison (i.e., *Please indicate whether the following sounds and/or sights bother you much more intensely than they do most other people*) in order to prompt responses that were more *atypical*, and therefore likely to trigger impairment/ distress.

*Frequency:* Participants were asked how often they were “bothered by a sound/sounds”. Although only 3.3% of participants reported being triggered more than 10 times per day, all response options were endorsed (see Table 3). Low endorsement rate of the higher frequency option was expected for a non-clinical sample. Therefore, it was decided that the response options remain unchanged.

**Table 3.** Endorsement Rates for Duke Misophonia Questionnaire Trigger Frequency Item.

| Response Option | Percent Endorsed |
| --- | --- |
| once a month or less | 21.7 |
| 2-3 times a month | 19.1 |
| 1-3 times a week | 22.4 |
| 4-7 times a week | 15.8 |
| 2-5 times per day | 13 |
| 6-10 times per day | 4.7 |
| > 10 times per day | 3.3 |

### Inter-scale Correlations

Table 4 shows the correlations among subscales of the DMQ. Subscale intercorrelations were all within the range of 0.430–0.839, indicating strong relationships between the proposed constructs. Based on the hypothesized structure of the subscales, the behavioral response scale was intended to represent a component of symptom severity along with affective, physiological and cognitive response. However, after reviewing the final items of the behavioral response subscale, it was discovered that this subscale conceptually overlapped more with the coping items. A symptom severity score representing the composite of all response items correlated at *r =* 0.991 with a severity score that did not include behavioral response, suggesting that the removal of the behavioral response items would not substantially affect scores on the overall symptom composite. Therefore, it was determined that behavioral response items be removed from the final scale.

**Table 4.** Correlations Among Subscale and Scale scores of the Duke Misophonia Questionnaire.

|  | 1 | 2 | 3 | 4 | 5 | 6 | 7 | 8 | 9 | 10 | 11 | 12 |
| --- | --- | --- | --- | --- | --- | --- | --- | --- | --- | --- | --- | --- |
| 1. Frequency of Response Item | 1 |  |  |  |  |  |  |  |  |  |  |  |
| 2. Affective Response Subscale | 0.601 | 1 |  |  |  |  |  |  |  |  |  |  |
| 3. Physiological Response Subscale | 0.460 | 0.658 | 1 |  |  |  |  |  |  |  |  |  |
| 4. Cognitive Response Subscale | 0.573 | 0.744 | 0.624 | 1 |  |  |  |  |  |  |  |  |
| 5. Behavioral Response Subscale<sup>a</sup> | 0.507 | 0.608 | 0.587 | 0.713 | 1 |  |  |  |  |  |  |  |
| 6. Coping-before Subscale | 0.561 | 0.603 | 0.553 | 0.719 | 0.724 | 1 |  |  |  |  |  |  |
| 7. Coping-during Subscale | 0.514 | 0.554 | 0.518 | 0.651 | 0.732 | 0.839 | 1 |  |  |  |  |  |
| 8. Coping-after subscale | 0.471 | 0.504 | 0.514 | 0.589 | 0.642 | 0.780 | 0.825 | 1 |  |  |  |  |
| 9. Impairment Scale | 0.430 | 0.525 | 0.469 | 0.601 | 0.535 | 0.572 | 0.530 | 0.581 | 1 |  |  |  |
| 10. Beliefs Scale | 0.545 | 0.626 | 0.621 | 0.706 | 0.621 | 0.69 | 0.652 | 0.614 | 0.664 | 1 |  |  |
| 11. Composite Symptoms Scale | 0.622 | 0.783<sup>b</sup> | 0.683<sup>b</sup> | 0.764<sup>b</sup> | 0.726 | 0.719 | 0.659 | 0.607 | 0.610 | 0.737 | 1 |  |
| 12. Composite Coping Scale | 0.55 | 0.592 | 0.562 | 0.698 | 0.753 | 0.853<sup>b</sup> | 0.882<sup>b</sup> | 0.840<sup>b</sup> | 0.590 | 0.696 | 0.707 | 1 |

*Note: Composite Symptoms Scale = composite score including affective, physiological and cognitive response items; Composite Coping Scale = composite score including coping-before, coping-during and coping-after items. <sup>a</sup>Behavioral response items were not included in the final scale; <sup>b</sup>corrected scale correlations represent correlations between the subscale and the composite score that does not include that subscale*

Consistent with the hypothesized structure, the affective, physiological and cognitive response scales correlated highly with the scale corrected symptom composite score (i.e., the composite score that did not include the subscale itself, with *r’*s ranging from 0.683 to 0.783). Also consistent with the hypothesized structure, high correlations were observed between the coping-before, coping-during and coping-after subscales and the scale corrected composite score (*r’*s ranging from 0.840 to 0.882).

### Composite Scale Analyses

#### Duke Misophonia Symptoms Scale

Combining the items from the affective response, physical response, and cognitive response subscales, the composite Symptoms subscale contained 23 items. Hierarchical item clustering indicated a four-cluster solution, with five sub-clusters interpreted as “Anger/Disgust” (five items, all from affect scale), “Anxiety/Panic” (12 items, three from affect scale, five from physical scale, and four from cognitive scale), “Aggression” (three items, all from cognitive scale), and “Escape” (three items, all from cognitive scale), although the anxiety/panic factor could be further subdivided into an affective/physical scale (eight items) and cognitive anxiety scale (four items). An exploratory bifactor model with five specific factors was then fit to the data, largely replicating the ICLUST solution. General factor saturation was strong (ω_H_ = 0.850), supporting the presence of an overarching “misophonia symptoms” factor and the utility of a symptoms composite score to operationalize this construct. The 23-item Symptoms scale was fit with a bifactor graded response model, with five specific factors derived from EFA factor loadings and the ICLUST results. This model fit the data well (*C*_2_(202) = 391.6, *p* < 0.001, CFI_C2_ = 0.990, RMSEA_C2_ = 0.048, SRMR = 0.043). Additionally, the resulting scale exhibited no local dependence (all LD-χ^2^ < 7.38), and no residual correlations greater than ±0.15. Marginal reliability for the general factor score was excellent (ρ_xx_ = 0.932). However, marginal reliability for the specific factors were relatively low (mean ρ_xx_ = 0.620, range [0.468, 0.781]), indicating that the Symptoms scale items are best interpreted as a single composite score rather than multiple subscale scores.

#### Duke Misophonia Coping Scale

Combining the items from the coping-before, coping-during, and coping-after subscales, the composite Coping scale contained 21 items. Hierarchical item clustering indicated a four-cluster solution, with clusters interpreted as “Cognitive Reframing” (three items, all from coping-during scale), “Sound Masking” (five items, three from coping-during scale and one each from coping-before and coping-after scales), “Self-soothing” (seven items, two from coping-before scale, one from coping-during scale, and four from coping-after scale), and “Distraction/Avoidance” (six items, two from coping-before scale and four from coping-during scale). An exploratory bifactor model with four specific factors was then fit to the data, largely replicating the ICLUST solution. General factor saturation was strong (ω_H_ = 0.841), supporting the presence of an overarching “misophonia coping strategies” factor and the utility of a symptom composite score to operationalize this construct. The 21-item Coping scale was fit with a bifactor graded response model, with four specific factors derived from the ICLUST/EFA results and three additional specific factors to account for dependencies between the items from the three coping timeframes (i.e., before, during, and after experiencing a trigger). This model fit the data well (*C*_2_ (145) = 262.2, *p* < 0.001, CFI_C2_ = 0.993, RMSEA_C2_ = 0.044, SRMR = 0.043). Additionally, the resulting scale exhibited no local dependence (all LD-χ^2^ < 9.59), and no residual correlations greater than ±0.15. Marginal reliability for the general factor score was excellent (ρ_xx_ = 0.929). However, with the exception of the “Masking” factor (ρ_xx_ = 0.846), marginal reliability for the specific factors were relatively low (mean ρ_xx_ = 0.492, range [0.320, 0.846]), indicating that the Coping scale items are best interpreted as a single composite score rather than multiple subscale scores.

### Convergent Validity Testing

#### Convergence Within Duke Misophonia Questionnaire

Strong correlations were observed between the Symptoms and Coping composite scales and between each of these with the Beliefs scale (*r*’s > 0.696, see Table 5). Additionally, large yet slightly lower correlations (*r*’s > 0.590) were observed between these three scales and the Impairment scale. The frequency of being bothered by a sound was more highly correlated with the Symptoms composite score (*r*_polyserial_ = 0.622) than the Impairment score (*r* = 0.430).

**Table 5.** Correlations between Duke Misophonia Questionniare and Existing Measures of Misophonia.

|  | 1 | 2 | 3 | 4 | 5 | 6 | 7 | 8 | 9 | 10 | 11 | 12 | 13 | 14 | 15 |
| --- | --- | --- | --- | --- | --- | --- | --- | --- | --- | --- | --- | --- | --- | --- | --- |
| DMQ Frequency of Response Item | 1 |  |  |  |  |  |  |  |  |  |  |  |  |  |  |
| DMQ Impairment Scale | 0.430 | 1 |  |  |  |  |  |  |  |  |  |  |  |  |  |
| DMQ Beliefs Scale | 0.545 | 0.664 | 1 |  |  |  |  |  |  |  |  |  |  |  |  |
| DMQ Composite Symptoms Scale | 0.622 | 0.610 | 0.737 | 1 |  |  |  |  |  |  |  |  |  |  |  |
| DMQ Composite Coping Scale | 0.550 | 0.590 | 0.696 | 0.707 | 1 |  |  |  |  |  |  |  |  |  |  |
| MQ Total | 0.673 | 0.598 | 0.717 | 0.822 | 0.710 | 1 |  |  |  |  |  |  |  |  |  |
| MQ Symptom Scale | 0.619 | 0.448 | 0.563 | 0.593 | 0.553 | 0.882 | 1 |  |  |  |  |  |  |  |  |
| MQ Emotions and Behaviors Scale | 0.596 | 0.611 | 0.717 | 0.865 | 0.712 | 0.924 | 0.635 | 1 |  |  |  |  |  |  |  |
| MQ Severity Scale | 0.641 | 0.579 | 0.643 | 0.669 | 0.656 | 0.756 | 0.636 | 0.724 | 1 |  |  |  |  |  |  |
| . MAQ Total | 0.534 | 0.672 | 0.841 | 0.721 | 0.607 | 0.748 | 0.591 | 0.747 | 0.694 | 1 |  |  |  |  |  |
| . MER Total | 0.559 | 0.516 | 0.675 | 0.806 | 0.627 | 0.799 | 0.572 | 0.845 | 0.632 | 0.717 | 1 |  |  |  |  |
| . MCR Total | 0.538 | 0.603 | 0.629 | 0.732 | 0.669 | 0.784 | 0.575 | 0.819 | 0.681 | 0.658 | 0.770 | 1 |  |  |  |
| . A-MISO-S Total | 0.646 | 0.618 | 0.708 | 0.731 | 0.676 | 0.783 | 0.612 | 0.787 | 0.801 | 0.740 | 0.748 | 0.722 | 1 |  |  |
| . MAS-I Emotional Response | 0.552 | 0.396 | 0.535 | 0.630 | 0.505 | 0.653 | 0.448 | 0.707 | 0.633 | 0.563 | 0.665 | 0.600 | 0.687 | 1 |  |
| . MAS-I Physical Response | 0.473 | 0.415 | 0.554 | 0.626 | 0.471 | 0.617 | 0.472 | 0.629 | 0.582 | 0.575 | 0.575 | 0.514 | 0.623 | 0.673 | 1 |

*note: DMQ = Duke Misophonia Questionnaire, MQ = Misophonia Questionnaire, MAQ = Misophonia Assessment Questionnaire, MER = Misophonia Emotional Responses Scale, MCR = Misophonia Coping Responses Survey, A-MISO-S = Amsterdam Misophonia Scale, MAS-I = Misophonia Activation Scale*

#### Convergence With Existing Misophonia Scales

Table 5 shows correlations between the composite and subscale scores of the DMQ and existing measures of misophonia. The Symptoms composite score of the DMQ is conceptually most similar to the Emotions and Behaviors Subscale of the MQ (Wu et al., 2014). Consistent with this, a high correlation of 0.865 supports convergent validity of the Symptoms composite scale. However, the Impairment score on the DMQ correlated 0.579 with the “Severity” item of the MQ, suggesting only moderate overlap. High correlations were observed (*r*’s ranging from 0.721 to 0.806) between scores of the MAQ, MER, MCR, and A-MISO-S and the Symptoms composite score of the DMQ. Slightly lower correlations of 0.626 to 0.630 were observed between the Symptoms composite score and the subscale scores of the MAS. Overall, the correlations between the DMQ Symptoms Scale and existing measures were higher than those observed between the Impairment scale and those same measures.

Next, analyses were conducted to assess the extent to which scores on the DMQ Symptoms Scale discriminated individuals with “clinical” and “sub-clinical” misophonia. First, relying on existing literature, clinical status was determined by scores on the MQ severity scale (i.e., < 7 suggests sub-clinical misophonia [*n* = 357]; ≥ 7 suggests clinical misophonia [*n* = 67]; Wu et al. 2014). Using the latent scores on the DMQ Symptoms scale, an ROC curve was constructed to predict misophonia classification. DMQ Symptoms scale scores demonstrated good ability to discriminate between those with clinical and sub-clinical misophonia (*AUC* = 0.82, 95% CI [0.77, 0.87]). Youden’s *J* index indicated an optimal cut point at a mean item score of 1.02 (on a 0–4 scale), resulting in a specificity and sensitivity of 0.72 and 0.78 respectively. Next, the criterion for determining clinical misophonia status was modified to include individuals with an average item score ≥ 2 on the MQ Emotions and Behaviors Scale and ≥ the MQ Severity Scale (*n* = 21). This was done for two reasons – (a) to increase the reliability of this criterion, which was likely limited when relying on only one item and (b) the authors believed it was conceptually meaningful to include the frequency of misophonic emotional and behavioral response in the criterion, in particular that a cut-off signifying at least “sometimes” experiencing all responses, was a conservative approach to delineating a clinical level of misophonia symptoms. Based on this, DMQ Symptoms scale scores demonstrated excellent ability to discriminate between clinical and sub-clinical misophonia (*AUC* = 0.97, 95% CI [0.95, 0.98]). Youden’s *J* index indicated an optimal mean score cut point of 1.8 (total score of 41.4), resulting in a specificity and sensitivity of 0.91 and 1 respectively.

#### Range for DMQ Impairment Scale scores

Specific ranges of scores on the severity scale of the MQ correspond to levels of symptom severity (minimal (1-3), mild (4-6), moderate (7-9), severe (10-12) and very severe (13-15; Wu et al., 2014). In order to aid in the interpretation of DMQ Impairment Scale scores, clinical ranges were derived using the percentile cut-points in the current sample, that corresponded to each clinical range on the MQ severity scale. Scores between 0-6 on MQ severity scale (corresponding to minimal to mild sensitivity) corresponded with a mean item score of 1.08 on the DMQ Impairment Scale (84.20% of participants scored in this range). The cut point of 10 on the MQ severity scale (suggesting predominantly moderate symptom level) corresponded with a mean item score of 3.17 on the DMQ Impairment scale (with 98.82% participants scoring up to this cut-point). Finally, a cut point of 15 on the MQ severity scale (indicating very severe sensitivity) corresponded to a mean item score of 4 on the DMQ Impairment Scale (i.e., 100^th^ percentile). Therefore, suggested clinical ranges for the DMQ Impairment scale are as follows: total score of 0-13 corresponds to “minimal-mild impairment” (cut point = 1.08 mean item score), total score of 14-38 corresponds to “moderate impairment” (cut point = 3.17 mean item score), total score of 39-48 corresponds to “severe to very severe impairment” (cut point = 4 mean item score).

## Discussion

The primary purpose of this study was to develop and psychometrically validate a self-report measure of misophonia. In addition to including items reflecting symptom severity and impairment in functioning, it was determined *a priori* to include items that include a wide spectrum of responses to misophonic triggers (affective, cognitive, physiological, behavioral), difficulties coping before, during and after being triggered, and dysfunctional beliefs related to misophonia. There were two phases of measure development. In Phase 1, items were generated and iteratively refined from a combination of the scientific literature and qualitative feedback from misophonia sufferers, their family members, and professional experts. In Phase 2, a large community sample of adults (*n* = 424) were recruited using the Amazon MTurk platform to complete DMQ candidate items and other measures needed for psychometric analyses. A series of iterative analytic procedures (e.g., factor analyses and IRT) were used to derive final DMQ items and subscales. From the overall item pool, the final DMQ (See Supplementary Table 1) is 86 items, and includes subscales: (1) Trigger frequency (16 items), (2) Affective Responses (5 items), (3) Physiological Responses (8 items), (4) Cognitive Responses (10 items), (5) Coping Before (6 items), (6) Coping During (10 items), (7) Coping After (5 items), (8) Impairment (12 items), and Beliefs (14 items). Composite scales were derived for overall Symptom Severity (combined Affective, Physiological, and Cognitive subscales) and Coping (combined the three Coping subscales). The analytic procedures used enable administration of the total DMQ, individual subscales, or the derived composite scales.

The DMQ is the first psychometrically validated measure of misophonia using an iterative item generation process with suggestions and feedback directly from individuals with high misophonia symptom severity and loved ones of those with misophonia. This grassroots approach to item generation was conducted to mitigate against any investigator biases or assumptions about misophonia that could otherwise have influenced the inclusion of items in the initial pool. Similarly, inclusion of these key stakeholders in the misophonia community during the item generation phase helped shape and refine the specific language used in the instructions and formatting of the DMQ, which may help with patient acceptability of the measure in clinical settings.

The DMQ is the first psychometrically validated measure of misophonia using factor analytic procedures combined with IRT in an English-speaking sample. IRT enables item-level analyses that can enhance the accuracy and reliability of a measure by mathematically discerning which items are the best fit with the underlying trait being assessed. As such, IRT provides an empirical basis for the inclusion of each item relative to other items and the underlying trait, supporting the internal validity of a scale. Although the MisoQuest is a promising new measure of misophonia and was developed using IRT (Siepsak et al., 2020), the item pool for this measure was constrained to reflect a proposed set of criteria that have not been empirically derived or conventionally accepted as definitive diagnostic criteria. Additionally, because it was validated with a Polish-speaking sample, the psychometric properties of the MisoQuest are unknown when used with English-speaking samples.

The DMQ is the first self-report measure of misophonia to include reliable and valid subscales reflecting a wide range of coping responses to misophonic triggers before, during, and after being exposed to these stimuli. Subscale intercorrelations indicated strong relationships between constructs. For example, the affective, physiological, and cognitive response scales correlated strongly with an overall symptoms composite score, whereas the coping before, during, and after subscales correlated highly with a coping scale composite score. Initially, we hypothesized that the behavioral response subscale would be correlated with overall symptom severity (along with the affective, physiological, and cognitive subscales). However, analyses indicated that it correlated more strongly with the coping subscale. Further, we discovered that removing the behavioral response subscale did not significantly alter overall scores. In an effort to reduce the overall length of the DMQ, the final measure omits items from the behavioral response subscale.

As hypothesized, the DMQ subscales were significantly positively correlated with other self-report measures of misophonia. More specifically, DMQ scores were associated with higher scores on the MQ (Wu et al., 2014), A-MISO-S (Schroder et al., 2013), Misophonia Activation Scale (MAS-I; Fitzmaurice, 2010), Misophonia Assessment Questionnaire (MAQ; Johnson, 2014), Misophonia Coping Responses Survey (MCRS; Johnson, 2014), and Misophonia Emotional Responses Scale (MERS; Dozier, 2015). The MisoQuest was not included in this study as it was not available at the time of data collection. Collectively, this pattern of results is evidence supporting the convergent validity of the DMQ.

The full DMQ may be used to assess symptoms, impairment in functioning, patterns of coping before, during, and after being triggered, and beliefs associated with misophonia. Alternatively, DMQ subscales and composite scales can be utilized to investigate changes in specific processes during treatment for misophonia. For example, the mechanisms of change in treatments targeting the use of coping skills can be evaluated using the DMQ across multiple times points before, during, and after treatment. Researchers or clinicians examining the effects of interventions on core beliefs and patterns of thinking in misophonia could use the DMQ Beliefs subscale as an endpoint. And, similarly, the impact of interventions on impairment in functioning in misophonia could be investigated using the DMQ Impairment subscale.

The DMQ Symptoms composite scale includes items reflecting anger, disgust, panic, and anxiety. Although other affective responses were less commonly observed, it is important to highlight that a minority of individuals with misophonia may endorse affective experiences beyond anger, disgust, panic, or anxiety. Nonetheless, these particular affective responses have been observed in a number of previous studies using other methodologies (e.g., Rouw & Erfanian, 2017). Similarly, the DMQ Symptoms scale includes items reflecting several specific physiological and cognitive responses to trigger cues, and, though these items were retained after careful iterative analyses, it should be noted that any given individual completing the DMQ may experience other responses not captured by the DMQ. Because the DMQ is limited to self-report assessment, it is recommended that a more comprehensive approach to misophonia include a functional analysis of common controlling variables influencing the probability of misophonic distress. Further, as suggested by others (Brout et al., 2018), it is recommended that such idiographic assessment be used with self-report measures as part of a broader multi-disciplinary assessment and treatment plan, including but limited to mental health providers (e.g., occupational therapy, audiology, neurology), that is personalized to the individual in context.

Analyses indicated that a cut point for clinical severity on the DMQ is a Symptoms composite scale mean item score of 1.8 or higher, which equates to a mean score of 41.4 or above. This was determined using the MQ (Wu et al., 2014) as a reference using Youden’s *J* Index. Similar processes were used to derive levels of impairment across DMQ Impairment subscale scores from minimal-mild (0-13), moderate (14-38), and severe (39-49). These cut points can be used until additional studies are done to replicate and cross validate the DMQ in other samples. Because there are no definitive diagnostic criteria for misophonia, it is advised that the cut point be used as a clinical indicator of possible caseness, not as a diagnosis per se. In addition, the DMQ cut point for severity should be used in the context of additional information about a given individual and the context in which they are experiencing misophonia symptoms. Individuals below the cut point may also be suffering significantly and, as such, it is not advised to make decisions that may impact individuals’ lives solely on the basis of the DMQ.

The results of this study should be considered within the context of its limitations. First, because our sample was drawn from MTurk and was not a clinical sample, some of the responses (e.g., physiological markers of hyperarousal) may have been endorsed at a lower rate than would be expected in a sample with a higher base rate and severity of misophonia. Future research cross validating the DMQ with clinical samples could specify, for example, which types of coping (e.g., cognitive reframing, sound masking, self-soothing, distraction, avoidance) are more or less adaptive amidst increased symptom severity. Evaluating the DMQ in both clinical and general population samples may lend insight into differential item functioning and phenotypic disparities on the various subscales. In general, exploration of the generalizability of the scale within large, more diverse samples is warranted.

Next, our analyses were conducted from cross-sectional data and we were thus limited in our ability to draw causal conclusions and establish predictive validity. Future longitudinal samples would allow for predictive analyses to test whether the frequency of response questions should be included in the severity scores along with the symptom scores. Exploring if and how strongly the frequency items predict clinical outcomes and impairment would determine their incremental validity to the existing symptom severity score. Longitudinal samples would also afford the opportunity to conduct test-retest reliability.

Furthermore, future validation studies should extend convergent validity analyses using additional misophonia measures that were not included in this current phenotypic study (e.g., MisoQuest, A-MISO-S). Another limitation in the current study was the relatively weak correlation between the MQ severity score and the DMQ Impairment scale score, perhaps because a one-item scale from the MQ inherently limits reliability. Including more misophonia measures will increase reliability and convergent validity of the DMQ. Although no semi-structured or structured interview measures of misophonia have been psychometrically validated, another important future direction will be to explore congruence between the DMQ and clinician-administered misophonia interviews. Additionally, the DMQ was developed with adults, yet misophonia commonly begins during childhood (Jager et al., 2020). As such, psychometrically validated measures of misophonia are needed for children under the age of 18. Lastly, distilling the DMQ into short and long forms in the future would increase clinical utility: a brief form would allow clinicians in both medical and psychiatric settings to readily screen for misophonia, and a long form would provide more comprehensive data as needed.

## Conclusions

The DMQ is the first psychometrically validated self-report measure of misophonia developed using a grassroots approach and multiple key stakeholders, iterative and rigorous analytic procedures to derive best fitting items (e.g., IRT), and a range of features commonly observed in misophonia beyond severity of symptoms and impairment in functioning. The DMQ can be used as a total score, with composite scores of symptom severity or difficulties coping, or subscales can be used individually. This is the first study demonstrating the psychometric properties of the DMQ. Accordingly, cross validation studies are needed to investigate the test-retest reliability and predictive validity of the DMQ across a range of samples.

## Data Availability

Data are available by request and per regulatory approval of the Duke University Medical Center IRB

## Notes

### Competing Interest Statement

Dr. Rosenthal is a member of the Scientific Advisory Board for the Misophonia Research Fund

### Funding Statement

Funding provided by anonymous philanthropic donors in support of the Duke Center for Misophonia and Emotion Regulation

### Author Declarations

Duke University Medical Center IRB Pro00101772 Development and validation of an adult self-report measure of Misophonia

## References

Arias, V. B., Garrido, L. E., Jenaro, C., Martínez-Molina, A., & Arias, B. (2020). A little garbage in, lots of garbage out: Assessing the impact of careless responding in personality survey data. Behavior Research Methods. https://doi.org/10.3758/s13428-020-01401-8

Beck AT. 1996. Beyond belief: A Theory of modes, personality, and psychopathology. In P. M. Salkovskis (Ed.), Frontiers of Cognitive Therapy, pp. 1–25. The Guilford Press.

Burns, G. L., Geiser, C., Servera, M., Becker, S. P., & Beauchaine, T. P. (2019). Application of the bifactor S–1 model to multisource ratings of ADHD/ODD symptoms: An Appropriate bifactor model for symptom ratings. Journal of Abnormal Child Psychology, 48, 881–894. https://doi.org/10.1007/s10802-019-00608-4

Cai, L. & Monro, S. (2014). A new statistic for evaluating item response theory models for ordinal data. National Center for Research on Evaluation, Standards, & Student Testing. Technical Report.

Cai, L., Yang, J. S., & Hansen, M. (2011). Generalized full-information item bifactor analysis. Psychological Methods, 16(3), 221–248. https://doi.org/10.1037/a0023350

Canu, W. H., Hartung, C. M., Stevens, A. E., & Lefler, E. K. (2016). Psychometric properties of the Weiss Functional Impairment Rating Scale: Evidence for utility in research, assessment, and treatment of ADHD in emerging adults. Journal of Attention Disorders, 1087054716661421. https://doi.org/10.1177/1087054716661421

Carpenter, K. L., Baranek, G. T., Copeland, W. E., Compton, S., Zucker, N., Dawson, G., & Egger, H. L. (2019). Sensory over-responsivity: an early risk factor for anxiety and behavioral challenges in young children. Journal of abnormal child psychology, 47(6), 1075–1088.

Cella, D., Riley, W., Stone, A., Rothrock, N., Reeve, B., Yount, S., … & Cook, K. (2010). The Patient-Reported Outcomes Measurement Information System (PROMIS) developed and tested its first wave of adult self-reported health outcome item banks: 2005–2008. Journal of Clinical Epidemiology, 63(11), 1179–1194. https://doi.org/10.1016/j.jclinepi.2010.04.011

Chandler, J., & Shapiro, D. (2016). Conducting clinical research using crowdsourced convenience samples. Annual Review of Clinical Psychology, 12, 53–81. doi:10.1146/annurev-clinpsy-021815-093623.

Chen, W. H., & Thissen, D. (1997). Local dependence indexes for item pairs using item response theory. Journal of Educational and Behavioral Statistics, 22(3), 265–289. https://doi.org/10.3102/10769986022003265

Clifton, J. D. W. (2020). Managing validity versus reliability trade-offs in scale-building decisions. Psychological Methods, 25(3), 259–270. https://doi.org/10.1037/met0000236

Dozier, T. H. (2015). Treating the initial physical reflex of misophonia with the neural repatterning technique: A Counterconditioning procedure. Psychological Thought, 8(2), 189–210. http://dx.doi.org/10.23668/psycharchives.1971

Dozier, T. H., Lopez, M., & Pearson, C. (2017). Proposed diagnostic criteria for misophonia: A multisensory conditioned aversive reflex disorder. Frontiers in psychology, 8, 1975.

Edelen, M. O., & Reeve, B. B. (2007). Applying item response theory (IRT) modeling to questionnaire development, evaluation, and refinement. Quality of Life Research, 16(1), 5. https://doi.org/10.1007/s11136-007-9198-0

Eid, M., Geiser, C., Koch, T., & Heene, M. (2017). Anomalous results in G-factor models: Explanations and alternatives. Psychological Methods, 22(3), 541–562. https://doi.org/10.1037/.met0000083.

Everaert, J., & Joormann, J. (2019). Emotion regulation difficulties related to depression and anxiety: A Network approach to model relations among symptoms, positive reappraisal, and repetitive negative thinking. Clinical Psychological Science, 7(6), 1304–1318. https://doi.org/10.1177/2167702619859342

Fackrell, K., Stratmann, L., Kennedy, V., MacDonald, C., Hodgson, H., Wray, N., … & Baguley, D. M. (2019). Identifying and prioritising unanswered research questions for people with hyperacusis: James Lind Alliance Hyperacusis Priority Setting Partnership. BMJ Open, 9(11). http://dx.doi.org/10.1136/bmjopen-2019-032178

Garcia-Garzon, E., Abad, F. J., & Garrido, L. E. (2019). Improving bi-factor exploratory modelling: Empirical target rotation based on loading differences. Methodology, 15(2), 45–55. doi:10.1027/1614-2241/a000163

Garcia-Garzon, E., Abad, F. J., & Garrido, L. E. (2020). On Omega Hierarchical Estimation: A Comparison of Exploratory Bi-Factor Analysis Algorithms. Multivariate Behavioral Research, 1-19. https://doi.org/10.1080/00273171.2020.1736977

Giordano, C., & Waller, N. G. (2019). Recovering bifactor models: A Comparison of seven methods. Psychological Methods, 25(2), 143–156. doi:10.1037/met0000227

Goetz, C., Coste, J., Lemetayer, F., Rat, A. C., Montel, S., Recchia, S., … & Guillemin, F. (2013). Item reduction based on rigorous methodological guidelines is necessary to maintain validity when shortening composite measurement scales. Journal of Clinical Epidemiology, 66(7), 710–718. https://doi.org/10.1016/j.jclinepi.2012.12.015

Golino, H. F., & Epskamp, S. (2017). Exploratory graph analysis: A New approach for estimating the number of dimensions in psychological research. PloS One, 12(6), e0174035. https://doi.org/10.1371/journal.pone.0174035

Golino, H., Shi, D., Christensen, A. P., Garrido, L. E., Nieto, M. D., Sadana, R., … & Martinez-Molina, A. (2020). Investigating the performance of exploratory graph analysis and traditional techniques to identify the number of latent factors: A Simulation and tutorial. Psychological Methods. 25(3), 292–320. https://doi.org/10.1037/met0000255

Goodman, W. K., Price, L. H., Rasmussen, S. A., Mazure, C., Fleischmann, R. L., Hill, C. L., et al. (1989). The Yale-Brown obsessive compulsive scale. I. Development, use, and reliability. Archives of General Psychiatry, 46(11), 1006–11.

Hauser, D., Paolacci, G., & Chandler, J. J. (2019). Common concerns with MTurk as a participant pool: Evidence and solutions. In F. R. Kardes, P. M. Herr, & N. Schwarz (Eds.), Handbook of Research Methods in Consumer Psychology (pp. 319–337). New York, NY: Routledge. doi: 10.31234/osf.io/uq45c

Kees, J., Berry, C., Burton, S., & Sheehan, K. (2017). An analysis of data quality: Professional panels, student subject pools, and Amazon’s Mechanical Turk. Journal of Advertising, 46(1), 141–155. https://doi.org/10.1080/00913367.2016.1269304

Kinnealey, M., & Oliver, B. (2002). Adult Sensory Questionnaire. Philadelphia, PA: Temple University, College of Allied Health Professionals.

Kinnealey, M., Oliver, B., & Willbarger, P. (1995). A phenomenological study of sensory defensiveness in adults. American Journal of Occupational Therapy, 49(5), 444– 451.

Lubbe, D. (2019). Parallel analysis with categorical variables: Impact of category probability proportions on dimensionality assessment accuracy. Psychological Methods, 24(3), 339–351. https://doi.org/10.1037/met0000171

Mackinnon, A., Jorm, A. F., Christensen, H., Korten, A. E., Jacomb, P. A., & Rodgers, B. (1999). A short form of the Positive and Negative Affect Schedule: Evaluation of factorial validity and invariance across demographic variables in a community sample. Personality and Individual Differences, 27(3), 405–416. https://doi.org/10.1016/S0191-8869(98)00251-7

Maydeu-Olivares, A., & Joe, H. (2014). Assessing approximate fit in categorical data analysis. Multivariate Behavioral Research, 49(4), 305–328. https://doi.org/10.1080/00273171.2014.911075

McKay, D., Kim, S. K., Mancusi, L., Storch, E. A., & Spankovich, C. (2018). Profile analysis of psychological symptoms associated with misophonia: a community sample. Behavior therapy, 49(2), 286–294.

McMahon, K., Anand, D., Morris-Jones, M., & Rosenthal, M. Z. (2019). A path from childhood sensory processing disorder to anxiety disorders: The mediating role of emotion dysregulation and adult sensory processing disorder symptoms. Frontiers in integrative neuroscience, 13, 22.

Miller, J. D., Crowe, M., Weiss, B., Maples-Keller, J. L., & Lynam, D. R. (2017). Using online, crowdsourcing platforms for data collection in personality disorder research: The Example of Amazon’s Mechanical Turk. *Personality Disorders: Theory*, Research, and Treatment, 8(1), 26–34. https://doi.org/10.1037/per0000191

Naylor, J., Caimino, C., Scutt, P., Hoare, D. J., & Baguley, D. M. (2020). The Prevalence and Severity of Misophonia in a UK Undergraduate Medical Student Population and Validation of the Amsterdam Misophonia Scale. Psychiatric Quarterly, 1-11.

Quek, T. C., Ho, C. S., Choo, C. C., Nguyen, L. H., Tran, B. X., & Ho, R. C. (2018). Misophonia in Singaporean psychiatric patients: a cross-sectional study. International journal of environmental research and public health, 15(7), 1410.

Revelle, W. (1979). Hierarchical cluster analysis and the internal structure of tests. Multivariate Behavioral Research, 14(1), 57–74. https://doi.org/10.1207/s15327906mbr1401_4

Revelle, W., & Condon, D. M. (2019). Reliability from α to ω A tutorial. Psychological Assessment, 31(12), 1395–1411. https://doi.org/10.1037/pas0000754

Rodriguez, A., Reise, S. P., & Haviland, M. G. (2016). Evaluating bifactor models: Calculating and interpreting statistical indices. Psychological Methods, 21(2), 137–150. https://doi.org/10.1037/met0000045

Rouw, R., & Erfanian, M. (2018). A large scale study of misophonia. Journal of clinical psychology, 74(3), 453–479.

Samejima, F. (1969). Estimation of latent ability using a response pattern of graded scores. Psychometrika Monograph Supplement, 34(4, Pt. 2), 100.

Siepsiak, M., Śliwerski, A., & Łukasz Dragan, W. (2020). Development and psychometric properties of misoquest—A new self-report questionnaire for misophonia. International journal of environmental research and public health, 17(5), 1797.

Siepsiak, M., Sobczak, A. M., Bohaterewicz, B., Cichocki, Ł., & Dragan, W. Ł. (2020). Prevalence of Misophonia and Correlates of Its Symptoms among Inpatients with Depression. International journal of environmental research and public health, 17(15), 5464.

Taylor, S. (2017). Misophonia: A new mental disorder?. Medical Hypotheses, 103, 109–117.

Toland, M. D., Sulis, I., Giambona, F., Porcu, M., & Campbell, J. M. (2017). Introduction to bifactor polytomous item response theory analysis. Journal of School Psychology, 60, 41–63. https://doi.org/10.1016/j.jsp.2016.11.001

Waller, N. G. (2018). Direct Schmid–Leiman transformations and rank-deficient loadings matrices. Psychometrika, 83(4), 858–870. doi:10.1007/s11336-017-9599-0

Watson, D., Clark, L. A., & Tellegen, A. (1988). Development and validation of brief measures of positive and negative affect: the PANAS scales. Journal of personality and social psychology, 54(6), 1063.

Watson, D., O’Hara, M. W., Simms, L. J., Kotov, R., Chmielewski, M., McDade-Montez, E. A., … & Stuart, S. (2007). Development and validation of the Inventory of Depression and Anxiety Symptoms (IDAS). Psychological Assessment, 19(3), 253–268. https://doi.org/10.1037/1040-3590.19.3.253

Wilcox, R. R., & Rousselet, G. A. (2018). A guide to robust statistical methods in neuroscience. Current Protocols in Neuroscience, 82(1), 8–42. https://doi.org/10.1002/cpns.41

Zhou, X., Wu, M. S., & Storch, E. A. (2017). Misophonia symptoms among Chinese university students: Incidence, associated impairment, and clinical correlates. Journal of Obsessive-Compulsive and Related Disorders, 14, 7–12.

