## Supplemental Table for "Development and Initial Validation of the Duke Misophonia Questionnaire"

Supplemental Table 1. All items generated for Duke Misophonia Questionnaire.

**Items**

**Affective Response Subscale**

I felt distressed.

I felt upset.

I felt guilty.

I felt scared.

**I felt hostile.**

I felt grouchy.

I felt ashamed.

I felt nervous.

**I felt jittery.**

I felt afraid.

**I felt anxious.**

I felt annoyed.

**I felt angry.**

**I felt disgusted.**

I felt unhappy.

**I felt frustrated.**

I felt rage.

I felt sad.

I felt disdain.

**I felt hateful.**

I felt embarrassed.

I felt devastated.

**I felt panic.**

I felt empathy for the person making the sounds.

I felt disengaged.

I felt detached.

I felt disconnected.

I felt empty.

I felt vacant.

My mind went blank.

I felt dazed.

**Physical Response Subscale**

I felt dizzy or lightheaded.

I had pain in my chest.

**I trembled or shuddered.**

**My heart pounded or raced.**

I felt nauseous.

I had an upset stomach.

I had trouble catching my breath.

I had hot spells.

I had cold spells.

I felt numbness or tingling in parts of my body.

I had a lump in my throat.

I felt physical pain.

I felt panicky.

I had goosebumps.

I felt my muscles tense up or tighten.

I started sweating.

I held my breath.

I had a headache.

I had groin sensitivity.

I clenched my teeth.

My ears hurt.

I flinched.

I felt like I was being prodded with a needle.

I felt a sensation similar to an electric shock.

I felt on edge.

I felt tension around my ears.

I had the urge to urinate.

My face got red.

**I became rigid or stiff.**

**I started breathing intensely or forcefully.**

I had a migraine.

I started tearing up.

**I reflexively jumped.**

**Cognitive Response Subscale**

**"I am helpless."**

"There is no hope."

"I resent the person making the sound."

"I wish that my senses would stop working."

"I am afraid I will hear that sound again."

"I want another person to know how upset I am."

"I want the person to stop making the sound."

"I want to force the person, thing or animal to stop making the sound."

"I want to mask the sound with something else."

"I want get physically far away from the sound."

"I am afraid I might hurt someone's feelings."

"I do not want to make a scene."

"I want to get away from the sound as quickly as possible, even if it would make a scene."

"I want to scream."

**"I want to cry."**

"I want to get revenge."

"I feel violated."

"This person is invading my space."

"They do not need to keep making this sound."

"Don't they know they are making this awful sound?"

"Don't they know how often they are making this awful sound?"

"Why can't they stop making this sound."

"They did not learn manners."

"They should not be making this sound."

"They do not care how this sound affects me."

"They know this sound hurts me and are making it on purpose."

"They are being disrespectful."

"They should stop making this sound."

"I am being irrational."

"Something is wrong with me."

"I am not normal."

"This is my fault."

"I hate the person making the sound."

"I am worried I might hurt the person making the sound."

**"How do I make this sound stop?"**

"Stop it!"

"I wish I could stop paying attention to the sound."

**"I need to get away from the sound."**

**"I would do anything to make it stop."**

"I hate this sound."

"Oh no! It's happening again!"

"Where is this sound coming from?"

"Why is this happening?"

"I would rather be dead than hear this."

"Who is making this sound?"

"They are going to do it again."

**"Everything is awful."**

"How will I live through this sound?"

"I can't escape."

"This is horrible."

"I can't believe they are doing this."

"I wish I could get over it."

"Why am I like this?"

"I am being attacked."

"I have to leave."

"How will I live my life like this?"

"They should know better."

"I need to make the sound stop."

"I should not be acting like this."

**"I cannot handle this."**

"This is not fair."

"This person is disgusting."

"Why can't they eat normally?"

**I thought about pushing, poking, shoving etc. the person making the sound.**

**I thought about screaming at, yelling at or telling off the person making the sound.**

**I thought about physically hurting the person making the sound.**

I imagined being violent toward the person making the sound but would never act on it.

**Behavioral Response Subscale**

I physically hurt someone.

I yelled at, screamed at, or told someone off.

I held myself back from physically hurting someone.

I held myself back from yelling at, screaming at, or telling someone off.

I nicely asked the person to stop making the sound.

I ordered the person to stop making the sound.

I discreetly covered one or both ears.

I calmly moved away from the noise.

I overtly covered my ears.

I mimicked the person making the sound.

I made repetitive sounds.

I turned away so that I did not see the person making the sound.

I pushed, poked, shoved etc. the person making the sound.

I immediately left the room to escape the sound.

I screamed.

I cried.

I physically hurt an animal.

I looked or stared at the person making the sound.

I showed my distress using my facial expression and/or body language (e.g., squinting my eyes, crossing my eyes, sighing loudly).

I blinked more.

I indirectly attempted to stop the sound (e.g., hand the person a tissue to get them to stop sniffling).

I took the sound producing object away from the person.

I kept asking the person to stop until they did.

I moved away from the sound.

I explained that the sound is bothersome to me.

I looked around for the sound source.

I purposefully tensed my muscles.

I held my breath.

I used ear plugs or headphones.

I rubbed my head.

I pulled my hair.

I tried to pop my ears.

I pushed one foot into the other.

I caused myself pain (e.g., dug my nails into my skin, bit my lip, pushed an object into my skin).

I showed how angry I was by acting aggressively (e.g., broke a pencil, slammed things).

I closed my eyes.

I blocked the sound source from my view (e.g., covered face with hair, covered eyes with hands, moved head so something else was in the way).

I made jerky movements.

I ran out of the room.

I produced an alternate sound (e.g., humming).

I grabbed my groin.

I clenched my fists.

I made repetitive movements (e.g., tapping, pacing).

**Coping-Before Subscale**

**I avoided certain people, places, or things so I would not have to hear sounds I dislike.**

I asked the person not to make a sound before they made it.

I did things to prevent a sound from occurring (e.g., asked person to take allergy pills, instituted a "no gum" policy).

I made it easy to escape if I needed to (e.g., sat near the door).

I positioned myself to limit sounds (e.g., moved computers away from my chair, sat facing away from a coworker).

I carried ear plugs, headphones or turned on ambient noise to block any sound that might occur.

I ate alone to avoid hearing bothersome sounds.

**I used a different sound to drown the bothersome sound (e.g., turned on TV).**

I determined where to live or work in order to avoid bothersome sounds.

**I used strategies to make myself less bothered by sounds I might hear (e.g., deep breathing, meditation, visualization).**

**I was on guard for bothersome sounds.**

I avoided looking at people or objects that might make a sound I dislike.

**I distracted myself so as not to be bothered by a sound I might hear.**

I held myself back from saying anything about a bothersome sound that might occur.

**I made a plan to cope with bothersome sounds if they occurred.**

I prepared myself to ignore the sound.

I isolated myself to avoid hearing a sound.

I described my sound sensitivity to others so they would understand.

I spent time learning about an event or activity to figure out if I would be bothered by the sounds there.

I brought soothing objects with me that would help me tolerate distress (e.g., soothing lotion, pictures).

I took medication to help me feel calmer.

I exercised to help me cope better.

I interacted with friends.

**Coping-During** **Subscale**

I asked the person to stop making the sound.

I left the room or situation to escape the sound.

I made another noise to cover the bothersome sound.

I posted on social media (e.g., Facebook) or an online community (e.g., Reddit) about the experience.

I asked for a break from the activity that put me in contact with the sound.

I alerted the person to the fact that they were making the sound.

I mimicked the person making the sound.

I used my facial expressions or body language to convey that I wanted the sound to stop (e.g., gave a side eye).

I texted or called someone else to help me cope with the ongoing sound.

I forced myself to tolerate the sound, i.e., "white knuckled it."

I acted like I was not bothered by the sound.

**I blocked the sound (e.g., covered ears with hands, headphones, ear plugs).**

**I increased the background noise to cover up the bothersome sound (e.g., turned on TV, rolled down car window).**

I moved to a different position.

I created a different physical sensation (e.g., dug nails into palm).

**I used strategies to calm myself (e.g., self-talk, breathing exercises).**

I deliberately focused my attention on the sound.

I tried to suppress thoughts about the sound.

**I changed my way of thinking about the sound.**

I took my anger out on an object.

I tried to soothe myself (e.g., ate comforting food, used a weighted blanket).

**I reminded myself that it could be worse.**

I removed the source of the sound.

**I looked away from the source of the sound.**

I indirectly asked the person to stop the sound (e.g., traded pens for one that does not click).

I asked a family member or friend to talk to others about the sound on my behalf.

I explained to the person that I have a condition that makes me sensitive to certain sounds.

I chewed at the same rate as the other person.

I reminded myself that the sound will end.

**I focused my attention on an activity (e.g., watched TV or videos).**

**I mindfully focused on current sensations without judgment.**

**I listened to music or a different sound.**

**I produced an alternate sound (e.g., humming).**

**Coping-After Subscale**

I told the person not to make the sound in the future.

I thought about what caused the sound.

I turned to others for emotional support.

I was nicer to the person making the sound than I had been before.

I told the person the effect the sound had on me.

I apologized for my reaction to the sound.

I told the other person about my sound sensitivity.

**I thought about strategies to help me cope better next time.**

I isolated myself from sources of unpleasant sounds.

**I did something to comfort myself (e.g., exercised, went somewhere calming, pet animals).**

I focused my attention on something else (e.g., watched TV, called a friend).

**I listened to a comforting sound (e.g., white noise, music).**

**I used the sight, smell or touch of an object to soothe myself (e.g., looked at a soothing picture, smelled a scent, or touched a soft blanket).**

I grounded myself in the present moment.

I laid down.

I sought out validation from social media (e.g., Facebook) or an online community (e.g., Reddit).

I held myself back from saying anything.

I wrote about my experience in a journal, blog, etc.

I asked the person why they made the sound.

I asked a family member or friend to talk to others on my behalf about the sound(s).

I discussed my concerns with a therapist or professional.

I went to sleep.

I suppressed thoughts about what happened.

I re-focused my attention on what I was doing before the sound occurred.

**I did some relaxation exercises (e.g., deep breathing, meditation).**

**Impairment Subscale**

My physical or mental health

My ability to interact in a group

**My ability to be with other people**

**My ability to function in daily activities without help**

How much time I spend with my family

The quality of my family relationships

Whether I feel like a good role model

**How much I enjoy spending time with my family**

What kind of future I imagine for my family

What kind of future I imagine for my long-term relationships

What I think about myself

**My self-esteem**

My confidence

How much I think other people like me

**How connected I feel to other people**

My productivity at work or school

**My performance at work or school**

**My ability to maintain employment**

My ability to gain employment

**My ability to work with others**

My ability to work without accommodations

My sleep

My day-to-day activities (e.g., meals, bathing, chores)

**The quality of my romantic relationships**

My desire to have a partner

My desire to seek out a partner

How much time I spend with my romantic partner

My ability to stay in a romantic relationship

How much time I spend with friends

What activities I'll do with friends

**The quality of relationships with my friends**

My ability to make new friends

My ability to keep friends

The kinds of activities I do with my family

The kinds of activities I do with my romantic partner

My finances (due to expenses related to my condition)

The relationships my family members have with each other (not including me)

**My ability to "be myself"**

My attendance at work or school

The kinds of people I am friends with

The number of friends that I have

**My ability to live with other people (e.g., roommate, partner)**

The kinds of activities I do by myself

**Beliefs Subscale**

"No one understands my problems with certain sounds."

"My sound issues do not seem to have a known cause."

"I am isolated."

"Others think I am crazy."

"No one can help me with my sound issues."

**"My sound issues will only get worse with time."**

"My sound issues have not been recognized as legitimate."

**"My whole life will be affected by sound issues."**

"I am not normal."

"It is difficult to explain what I am going through."

**"People do not understand me."**

"Nobody loves me."

"I am not a whole person."

"Nobody will ever love me."

**"I am a burden on others."**

"No one believes me."

**"No one can help me."**

**"I hate being like this."**

"I am being too difficult."

"No one cares about me."

"I am not like others."

"People do not understand my experience."

"Others are inconsiderate."

**"This is unfair."**

"I am missing out on a lot."

"I am a bad person."

"People think I am crazy."

**"I am crazy."**

"I am not happy."

"This consumes my attention."

**"I should get over it."**

"This is ridiculous."

**"I should have known how to cope earlier."**

"I have suffered."

"This will affect my long-term relationships."

**"I am weak."**

"I have a disability."

"I need to work harder than others to make up for this problem."

"I need to control this experience."

**I should be able to control my reaction to these sounds**

**"My reactions to sounds are irrational."**

**"I will be rejected if people find out."**

"I am a hypocrite as I make similar sounds myself."
